# Exploring the relationship between knowledge, attitudes, and ENDS use among youth in Pakistan: Implications for effective tobacco control strategies

**DOI:** 10.1101/2025.04.13.25325758

**Authors:** Madeeha Malik, Azhar Hussain, Ayisha Hashmi

## Abstract

Tobacco use remains a major public health issue, especially among youth, with the rising use of Electronic Nicotine Delivery Systems (ENDS) posing new challenges for tobacco control. This study explores the relationship between knowledge, attitudes, and ENDS use among youth in Pakistan, focusing on socio-demographic and behavioral factors influencing smoking behavior. An exploratory cross-sectional study was conducted using a Computer-Assisted Personal Interviewing (CAPI) survey across 13 major cities in Pakistan. Participants aged 15–24 years were selected through convenience sampling, yielding a final sample of 462 respondents. A structured, pilot-tested questionnaire (Cronbach’s α = 0.76) assessed demographics, ENDS knowledge, attitudes, and smoking behaviors. Data was analyzed using SPSS Version 26 with descriptive statistics, Chi-square tests, Spearman’s correlation, and multivariate logistic regression. Among respondents, 83.5% were male, 30.7% aged 18–20 years, 59.7% resided in urban areas, and 71.2% were students. Significant knowledge gaps were found, with many youth incorrectly believing that ENDS are nicotine-free or less harmful than cigarettes. Parental monitoring and student status were protective factors. Urban residence (AOR = 1.44, p = 0.0125) and male gender (AOR = 5.22, p = 0.009) were significant predictors of ENDS use, while employment (AOR = 0.43, p = 0.0146) and physical activity (AOR = 0.66, p = 0.0004) were associated with less favorable attitudes toward ENDS. Knowledge alone does not deter ENDS use. Social influences and attitudes significantly shape behavior, highlighting the need for targeted interventions addressing peer influence, misinformation, and stronger regulatory measures.

## Introduction

The rising use of Electronic Nicotine Delivery Systems (ENDS) among youth has raised worldwide public health concerns. ENDS, marketed initially as a harm reduction alternative to combustible tobacco, have grown in use, especially among adolescents and young adults ^[1]^. Yet their swift uptake has prompted important questions about knowledge, attitudes, and socio-demographic factors associated with ENDS use in youth ^[2]^. Youth perceptions regarding ENDS are recognized as varied based on potential misunderstanding, targeted capital campaigns, and social standards ^[3]^. Furthermore, socio-demographic characteristics, including age, sex, educational background, socioeconomic status, and family background, can also adequately predict attitudes towards ENDS ^[4]^. Awareness of ENDS and their potential harms affects smoking behavior considerably. Research shows that many youths believe that these products are less harmful than traditional cigarettes, which may explain their pervasiveness ^[5]^. Willett et al. (2019) reported that almost half of adolescent ENDS users did not know that nicotine was present in E cigarettes, representing major gaps in awareness about the health effects of ENDS ^[4]^. According to the Global Youth Tobacco Survey ^[6]^, over 30% of young ENDS users erroneously believe that there is no risk involved with vaping at all, supporting the need for comprehensive public health education. According to a study by Islam et al. (2022) young people who believe that ENDSs are "cool," "modern," or "socially acceptable" are significantly more likely to try vaping ^[7]^. This finding mirrors that of Cullen (2018) who found that social desirability and peer pressure are driving users to persist with their use of ENDS despite knowledge of the potential health risks ^[2]^.

Pakistan has an estimated 22 million adult smokers, which causes a huge burden on smoking related diseases (lung cancer, chronic obstructive pulmonary diseases (COPD), and cardiovascular diseases) ^[8]^. While the government has implemented measures to control tobacco consumption through taxation policies, health warnings on cigarette packets, and the prohibition of smoking in public spaces, the implementation mechanism is weak, and the market is largely informal and unregulated. The emergence of ENDS has added an additional layer of complexity since these products continue to be insufficiently regulated. Unlike combustible cigarettes, ENDS are widely available from online retailers and vape shops, often with minimal to no restrictions on sales, and their use has been associated with increased accessibility among youth. The popularity of flavored E-cigarettes as affordable, easily accessible alternatives has also played a significant role in their proliferation, especially in urban centers like Karachi, Lahore, and Islamabad with vape lounges and international chains of specialty stores that sell these products purporting to be lifestyle accessories rather than tobacco cessation products. Such uncontrolled expansion is a clear call for implementing evidence-based policies that seek to balance harm reduction for current smokers while preventing nicotine addiction in the non-smoking population.

In Pakistan, the absence of explicit guidelines on the regulation of ENDS has enabled the tobacco industries to flourish with little government scrutiny. As cigarette manufacturers contend with taxes and advertising restrictions, ENDS companies continue to operate in a legal gray area and promote themselves to youth via social media marketing and influencer-sponsored content. This trend reflects trends in other developing countries, where poorly defined regulatory structures enable ENDS companies to exploit nascent markets. Furthermore, public knowledge and attitudes towards the use of ENDS in Pakistan are variable and often driven by misinformation. Indoor air quality can be a distinct but equally prominent vector in this battle, as many young people believe that vaping is either totally harmless or much less dangerous than traditional cigarettes, likely due to misleading marketing and the lack of effective public health messaging ^[9]^. Awareness about the risks of cigarette smoking is fairly high, but knowledge about the potential harms of ENDS, nicotine dependence, and regulatory policies is low. Moreover, perceptions of ENDS need to be interpreted with caution; for some youth, ENDS as a phenomenon seem trendy and socially acceptable compared to smoking, while for others, ENDS are perceived as a potential tool for quitting cigarettes. According to research, urban youth are more likely to have positive perceptions of ENDS, attributed to higher exposure to social media promotions about these products and exposures to established vape culture, while rural populations score low on awareness and usage ^[10]^. However, there is increasing consensus among public health experts that ENDS may help some people quit smoking, especially in a country to which more conventional nicotine replacement therapies (NRTs) including nicotine gums and patches are still largely unexplored. Research by McNeill et al. (2020) concluded that if effectively controlled, ENDS may help current smokers reduce or quit the use of combustible tobacco, a key issue for Pakistan, due to its high adult smoking population ^[1]^. Yet in the absence of sensible regulation, ENDS risk becoming another pathway to nicotine dependence for nonsmokers, particularly young people. With the rising utilization of ENDS in Pakistan, there has been little research measuring knowledge and attitudes towards these products, especially among the youth. This will not only continue to be relevant as users shift from traditional cigarette smoking to ENDS but it also demonstrates an area of research not heavily covered in existing literature.

Understanding youth’s knowledge and attitudes about ENDS could help healthcare professionals determine how likely a youth is to smoke, which can help guide efforts to promote initiatives targeting the initiation of use of nicotine products and smoking cessation among youth. Individual, social and contextual factors are associated with youth use of E-cigarettes. Exploring predictors of ENDS use among youth and delineating the dynamics of public awareness, risk perceptions and behavioral patterns concerning ENDS is pertinent for shaping evidence-based policy and risk-reduction interventions in the country. Therefore, the present study was designed to examine the relationship between socio-economic, health, and behavioral factors with knowledge and attitudes toward ENDS use among youth. By identifying key determinants influencing perceptions and usage patterns, this research seeks to provide evidence-based insights to guide public health policies, awareness campaigns, and harm reduction strategies related to ENDS use in Pakistan.

## Methodology

### Study Design

This study adopted an exploratory cross-sectional design using a Computer-Assisted Personal Interviewing (CAPI) survey to assess and compare knowledge and attitudes toward ENDS among youth in Pakistan. The study also examined the influence of various socio-economic, health, and behavioral factors associated with ENDS use.

### Ethical Considerations

Ethical approval for this study was obtained from the Ethical Review Committee (ERC) of Pak-Austria Fachhochschule University, Haripur under ERC Application Number: PAF-IAST/2024/27. The study was conducted in strict accordance with the ethical principles outlined in the Declaration of Helsinki, ensuring the protection, dignity, and rights of all participants. Before participation, informed written consent was obtained from all the respondents, outlining the study’s objectives, potential risks, and confidentiality measures. For participants under the age of 18, additional parental or guardian consent was received. Confidentiality agreements were established to protect personal information, and all data was anonymized and securely stored to prevent unauthorized access. Participants were informed of their right to withdraw from the study at any stage without any consequences. The study-maintained transparency, voluntary participation, and non-coercion to uphold ethical research standards. Data collected were strictly used for research purposes only, ensuring compliance with international ethical guidelines for human subject research.

### Inclusion and Exclusion Criteria

This study targeted youth aged 15 to 24 years, following the World Health Organization (WHO) definition, which classifies youth as “individuals within this age range (15 – 24 years), distinct from adolescents (10–19 years) and young adults (20–24 years)”. To ensure relevance, participants were required to be residents of one of the 13 selected district cities in Pakistan, able to read and understand English or Urdu, and willing to provide informed written consent (with parental or guardian consent for those under 18). Individuals younger than 15 or older than 24 years, those with language barriers, those unwilling to participate, and those with substantial missing or incomplete survey responses were excluded from the study. This inclusion and exclusion process ensured that data were collected from a diverse and representative sample of Pakistani youth, providing valuable insights into their knowledge, attitudes, and behaviors toward ENDS use.

### Sampling Strategy and Sample Size Calculation

The sample size for this study was determined using statistical methods to ensure representative and generalizable findings regarding youth knowledge, attitudes, and behaviors toward ENDS use. Based on the 2017 Census of Pakistan, the total youth population (15–24 years) was 39,862,513, and the sample size was calculated using a 95% confidence interval, a 5% margin of error, and a response distribution of 50%. The sample size was derived using Cochran’s formula, given as:

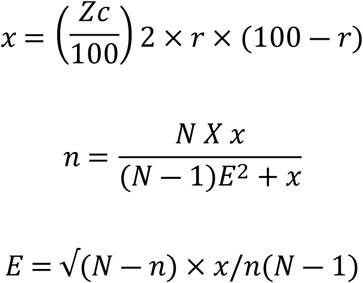

where:

- Z = Z-score for a 95% confidence level (1.96)
- r = Estimated proportion of the population with the characteristic of interest (50% or 0.5)
- N = Total youth population (39,862,513)
- E = Margin of error (5% or 0.05)

Using this formula, the initial sample size was calculated to be 385 respondents, which was then adjusted to 462 respondents to account for a 20% anticipated dropout rate, ensuring statistical robustness.

A convenience sampling technique was used, selecting youth from 13 major district cities across Pakistan, ensuring geographic, demographic, and socio-economic diversity. These cities included Lahore, Faisalabad, Rawalpindi & Bahawalpur (Punjab); Karachi, Nawabshah & Larkana (Sindh); Peshawar, Haripur (Khyber Pakhtunkhwa); Quetta (Balochistan); Muzaffarabad (Azad Jammu & Kashmir); Gilgit (Gilgit-Baltistan); and Islamabad Capital Territory (ICT). This represents an inclusive approach to the recruitment of youth participants, particularly those based in urban and semi-urban settings, who are increasingly susceptible to the influence of changing social norms, tobacco marketing, and access to ENDS.

### Development of Study Tool

To ensure the reliability and validity of the study, a structured survey questionnaire was developed to assess knowledge, attitudes, and factors influencing ENDS use among youth in Pakistan. The tool was designed based on an extensive literature review of prior studies on ENDS, smoking behavior, and tobacco control policies. The survey instrument was structured to collect demographic, socio-economic, and behavioral data, along with specific questions evaluating respondents’ knowledge and perceptions of ENDS, smoking history, and influences shaping their attitudes and behaviors.

To enhance the content validity of the questionnaire, a focus group discussion (FGD) was conducted with a panel of experts in the fields of tobacco control, alternative nicotine delivery systems (ANDS), and public health research. The expert panel reviewed the relevance, clarity, and comprehensiveness of the questions, ensuring that the instrument effectively measured the intended variables. Additionally, the face validity of the tool was evaluated by pre-testing the questionnaire among a small sample of youth to assess its readability, comprehension, and response time.

A pilot study was conducted on 10% of the total sample size (approximately 46 respondents) to test the feasibility and effectiveness of the tool. The pilot testing helped identify ambiguous or difficult-to-understand questions, which were refined based on participant feedback. The internal consistency and reliability of the questionnaire were measured using Cronbach’s Alpha (α), which came to be 0.76 and was considered acceptable for ensuring reliability. Furthermore, construct validity was assessed using Principal Component Analysis (PCA) to confirm that the questionnaire effectively captured the constructs of knowledge, attitudes, and behavioral factors related to ENDS use.

The final questionnaire was divided into multiple sections: Section 1: Demographic Information Age, gender, education level, employment status, household income, and place of residence; Section 2: Knowledge of ENDS – Awareness of nicotine content, health risks, regulatory policies, and differences between ENDS and traditional cigarettes; Section 3: Attitudes Toward ENDS – Perceptions regarding safety, social acceptability, harm reduction, and ENDS as a smoking cessation tool and; Section 4: Behavioral and Influencing Factors – History of smoking, exposure to tobacco advertisements, peer influence, and parental monitoring. To facilitate data collection, the questionnaire was digitized and integrated into the Zoho Survey Pro platform, allowing responses to be recorded electronically using internet-enabled tablets. The Computer-Assisted Personal Interviewing (CAPI) technique was employed, where trained data collectors administered the questionnaire, read the questions aloud, and recorded participant responses in real time. The finalized and validated survey tool ensured that data collection was comprehensive, reliable, and aligned with the study objectives, providing meaningful insights into youth knowledge, attitudes, and behaviors related to ENDS use in Pakistan.

### Data Collection

The data collection process was carefully designed to ensure accuracy, reliability, and ethical compliance while gathering information on youth knowledge, attitudes, and behaviors toward ENDS use. The study utilized a Computer-Assisted Personal Interviewing (CAPI) survey method, where trained data collectors administered the questionnaire using internet-enabled tablets equipped with the Zoho Survey Pro platform. The CAPI approach allowed real-time data entry, minimizing errors associated with manual recording while ensuring structured and standardized responses.

### Recruitment and Training of Data Collectors

Before initiating data collection, a team of trained field interviewers was recruited to conduct face-to-face interviews. All data collectors underwent an intensive training program, which included:

- Understanding the study objectives and methodology
- Following ethical guidelines and informed consent procedures
- Training on the CAPI technique and Zoho Survey Pro platform
- Ensuring proper communication with participants
- Addressing participant concerns and clarifying questions
- Handling situations where participants declined consent or withdrew from the study

The training also covered professional ethics, confidentiality protocols, and participant rights, ensuring that the data collection process adhered to ethical standards.

### Participant Recruitment and Consent Process

Participants were recruited from 13 major district cities across Pakistan, ensuring diverse socio-economic and geographic representation. The study targeted youth aged 15-24 years from urban and semi-urban populations. To ensure voluntary participation, informed written consent was obtained from all respondents before administering the questionnaire. For participants under 18 years of age, parental or guardian consent was also secured. Participants were informed about the study’s purpose, confidentiality measures, and their right to withdraw at any time without consequences.

### Survey Administration and Data Entry

The structured questionnaire was administered in person using the CAPI technique. The data collectors read each question aloud to the participant, provided explanations when necessary, and recorded responses in real time. The Zoho Survey Pro platform allowed for automatic data entry, reducing risks of missing or incorrect responses. To ensure data completeness and quality, built-in validation checks were included in the Zoho platform, preventing skipped or inconsistent responses. Data collectors also conducted spot checks to verify accuracy before submitting the responses.

### Data Security and Confidentiality

All collected data were anonymized to protect participant identities and securely stored on password-protected servers. Only authorized research personnel had access to the data, ensuring compliance with ethical and confidentiality standards.

### Quality Control Measures

To maintain data accuracy and reliability, the following quality control measures were implemented:

- Pilot Testing: The questionnaire was pre-tested on 10% of the total sample size (46 participants) to identify and correct potential issues before full-scale data collection.
- Supervisory Review: Field supervisors monitored data collectors and conducted random back-check interviews with respondents to verify responses.
- Daily Data Audits: Collected data were reviewed daily for completeness, logical consistency, and outliers.
- Real-Time Monitoring: The Zoho platform enabled real-time tracking of data collection progress, ensuring that sampling quotas were met across all target cities.

### Challenges and Mitigation Strategies

Several challenges were anticipated during data collection, including participant hesitancy, misinterpretation of questions, and access to respondents in different geographic locations. To address these:

- Data collectors were trained in effective communication techniques to gain participant trust.
- Survey questions were tested and refined for clarity and cultural appropriateness.
- The study was conducted in public spaces, educational institutions, and youth centers, ensuring accessibility to respondents

### Data Analysis

After completing the data collection process, the dataset was carefully cleaned, validated, and analyzed using IBM SPSS Version 26. The data analysis involved descriptive, inferential, and multivariate statistical techniques to examine the relationships between knowledge, attitudes, and behavioral factors influencing ENDS use among youth in Pakistan. Since knowledge and attitude scores were not normally distributed, Spearman’s rank correlation was used to examine the association between knowledge and attitude toward ENDS. The correlation coefficient (ρ, rho) measured the strength and direction of this relationship, with statistical significance set at p < 0.05. While Chi-square test was used to analyze the associations between various factors and smoking behavior. To identify the key predictors of ENDS knowledge and attitudes, multivariate logistic regression analysis was performed. The model adjusted for potential confounders and calculated adjusted odds ratios (AORs) with 95% confidence intervals (CIs) to assess the independent effects of demographic, socio-economic, and behavioral factors on knowledge and attitudes toward ENDS. A p-value < 0.05 was considered statistically significant in all analyses.

## Results

### Demographics Characteristics of Respondents

Of the 462 participants surveyed, 83.5% (n=386) were male and 16.5% (n=76) were female. The majority were aged 18-20 years (30.7%, n=142) and 23-25 years (30.1%, n=139). Urban residents comprised 59.7% (n=276), while 40.3% (n=186) lived in rural areas. Punjab had the highest representation (31.0%, n=143), followed by Sindh (22.7%, n=105) and Khyber Pakhtunkhwa (15.4%, n=71). Most participants held a bachelor’s degree (46.5%, n=215), and 18.2% (n=84) completed a master’s degree. Students made up 71.2% (n=329), with 17.1% (n=79) engaged in part-time and 9.5% (n=44) in full-time employment. Regarding economic status, 34.0% (n=157) of households had a monthly income of PKR 100,000–199,999, while 20.6% (n=95) earned PKR 50,000–99,999. Most participants (71.2%, n=329) had no personal income, and 25.8% (n=119) did not receive pocket money. Islam was the predominant religion (99.6%, n=460). About 75.5% (n=349) lived with parents/guardians, and 10.6% (n=49) in hostels. Family structures were nearly evenly split between joint (50.4%, n=233) and separate (49.6%, n=229) households.

In terms of health and lifestyle, 95.7% (n=442) reported no diagnosed diseases. Daily physical activity was reported by 27.9% (n=129), while 6.3% (n=29) never exercised. Alcohol use was low (4.5%, n=21), and 93.5% (n=432) reported no illicit drug use. Small percentages reported using marijuana (1.3%, n=6), methamphetamine (1.5%, n=7), and prescription drugs for non-medical purposes (0.2%, n=1). Parental education was high (69.9%, n=323), with 35.3% (n=163) parental monitoring. A detailed description is given (Table 1).

**Table 1:**
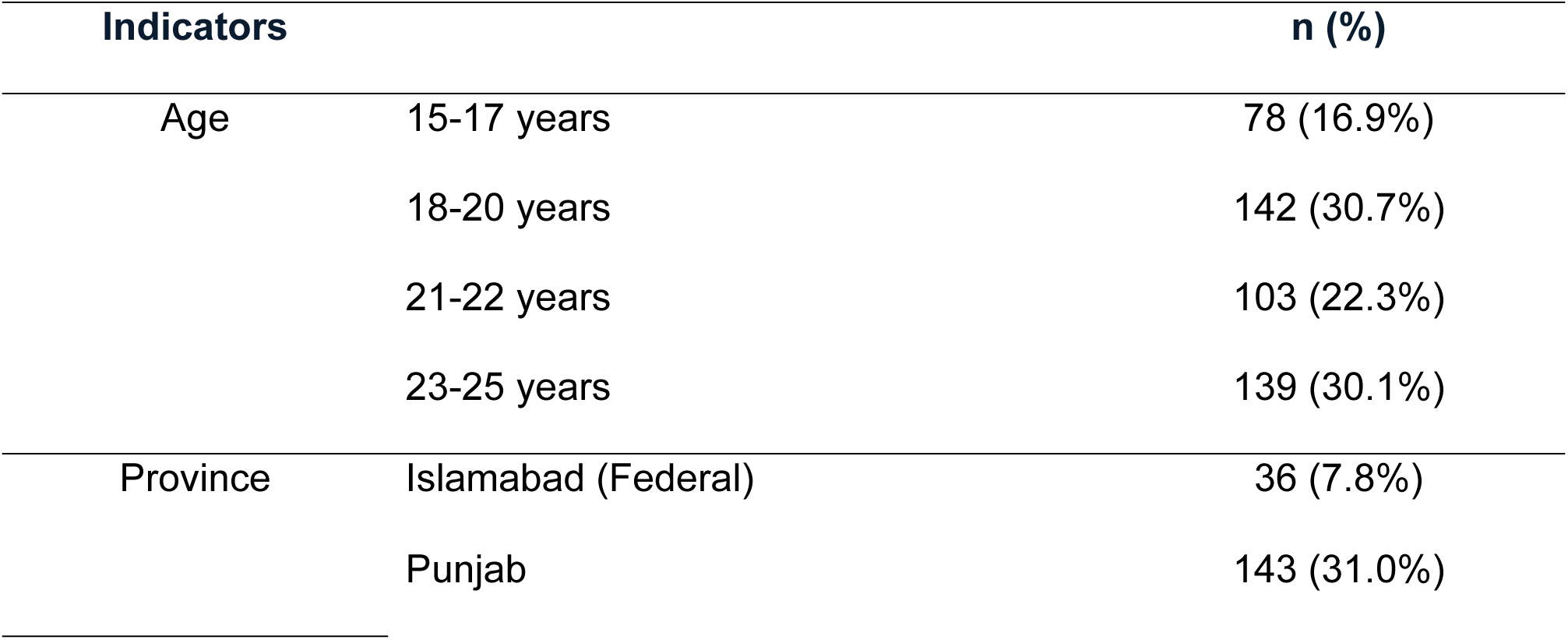
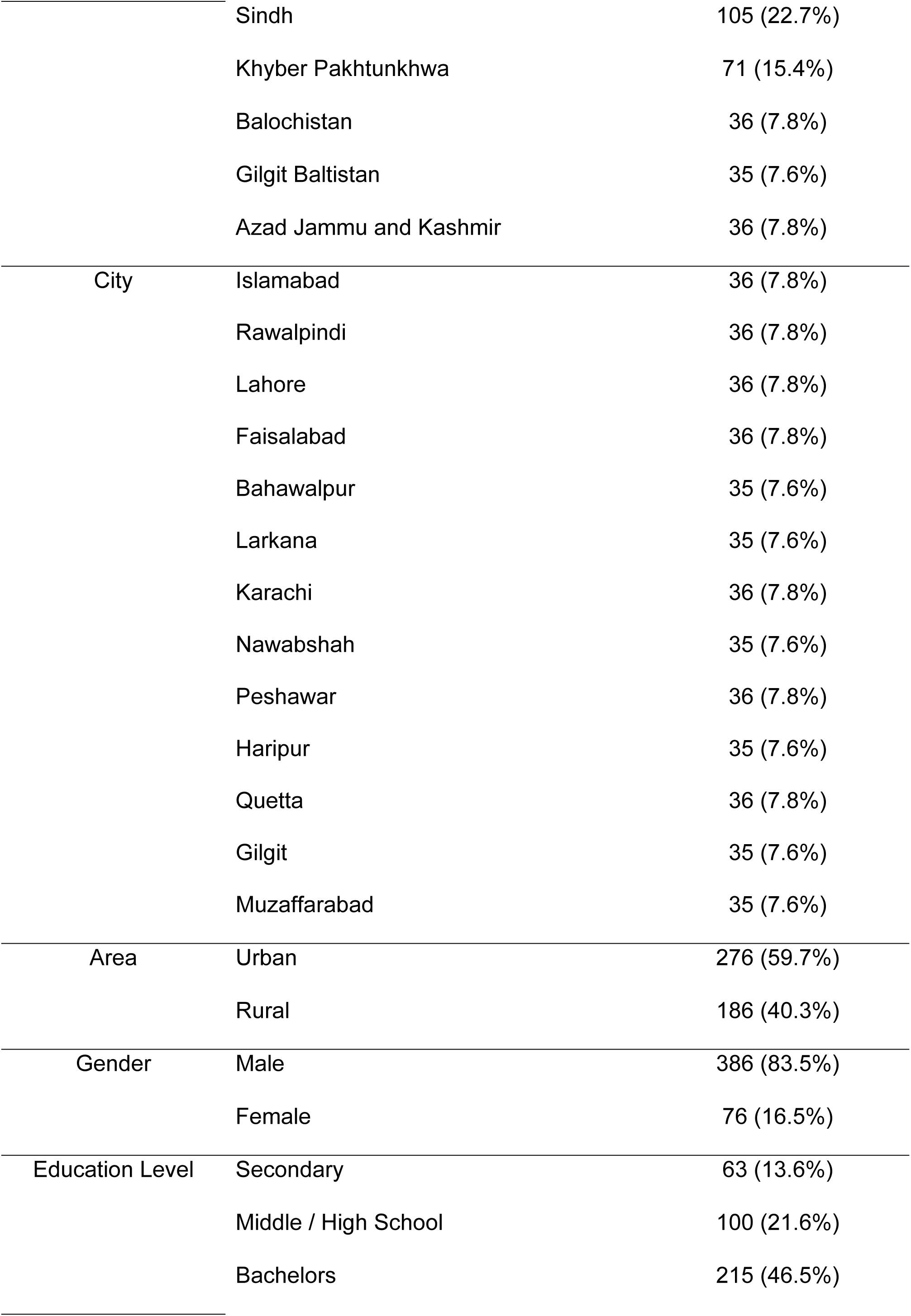
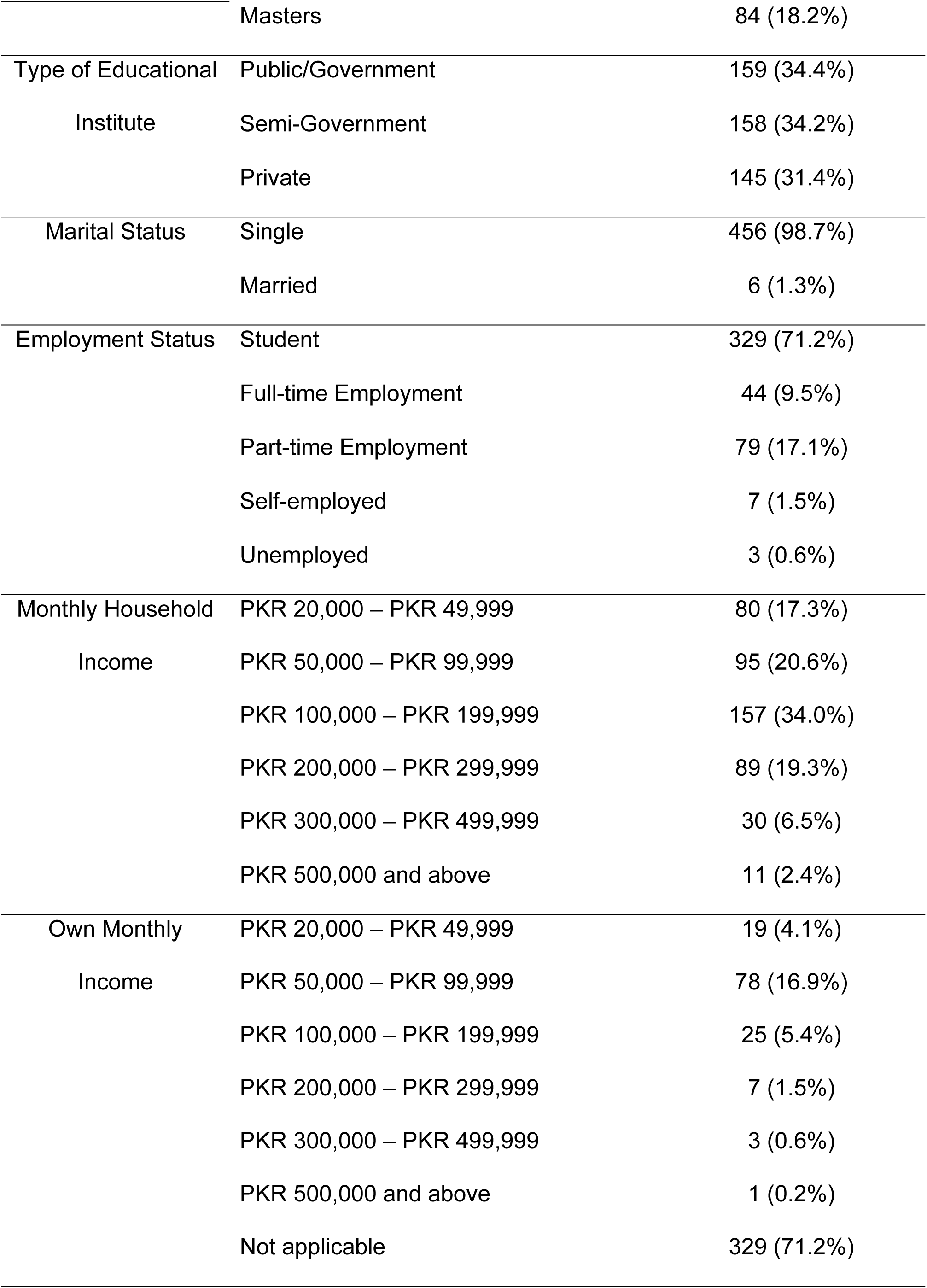
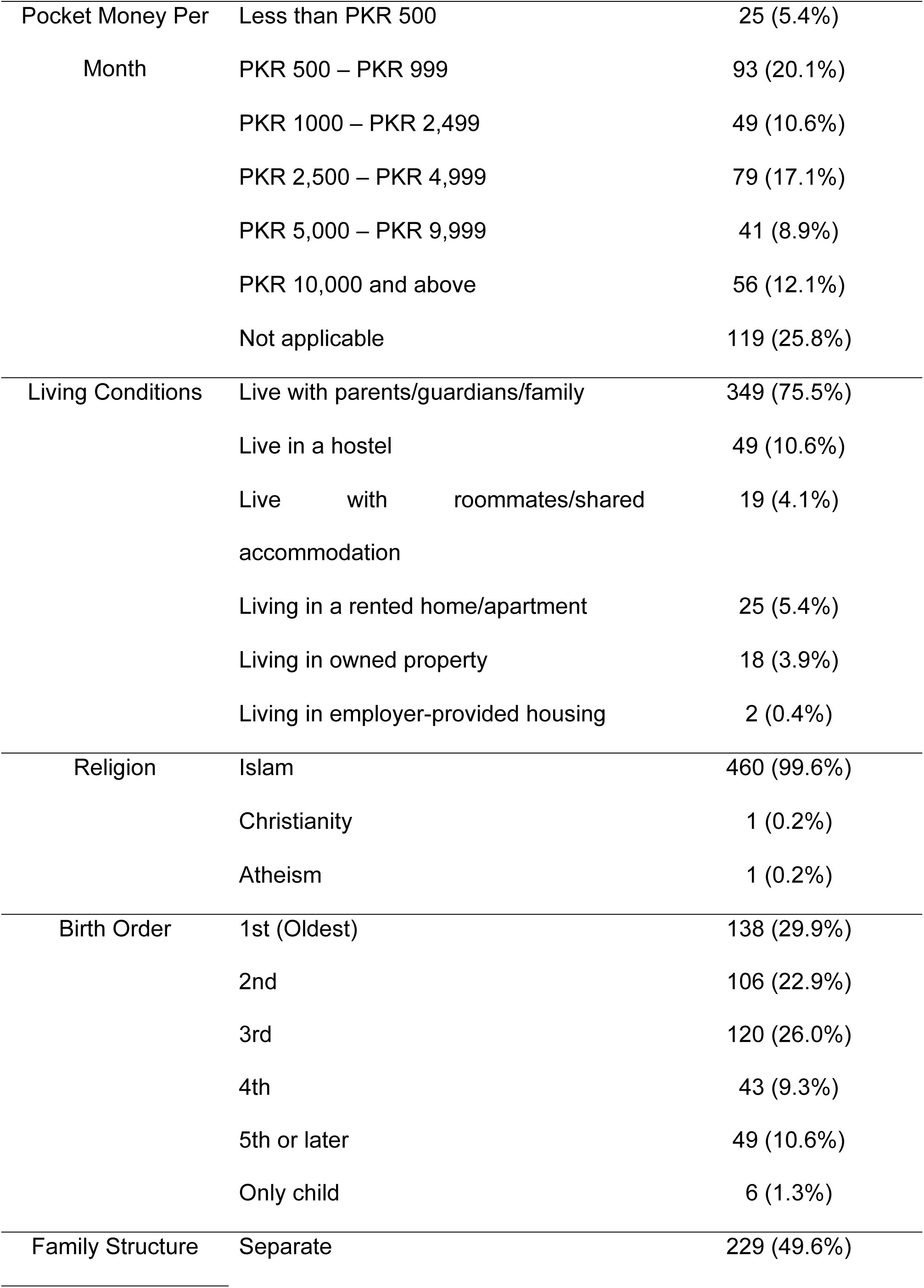
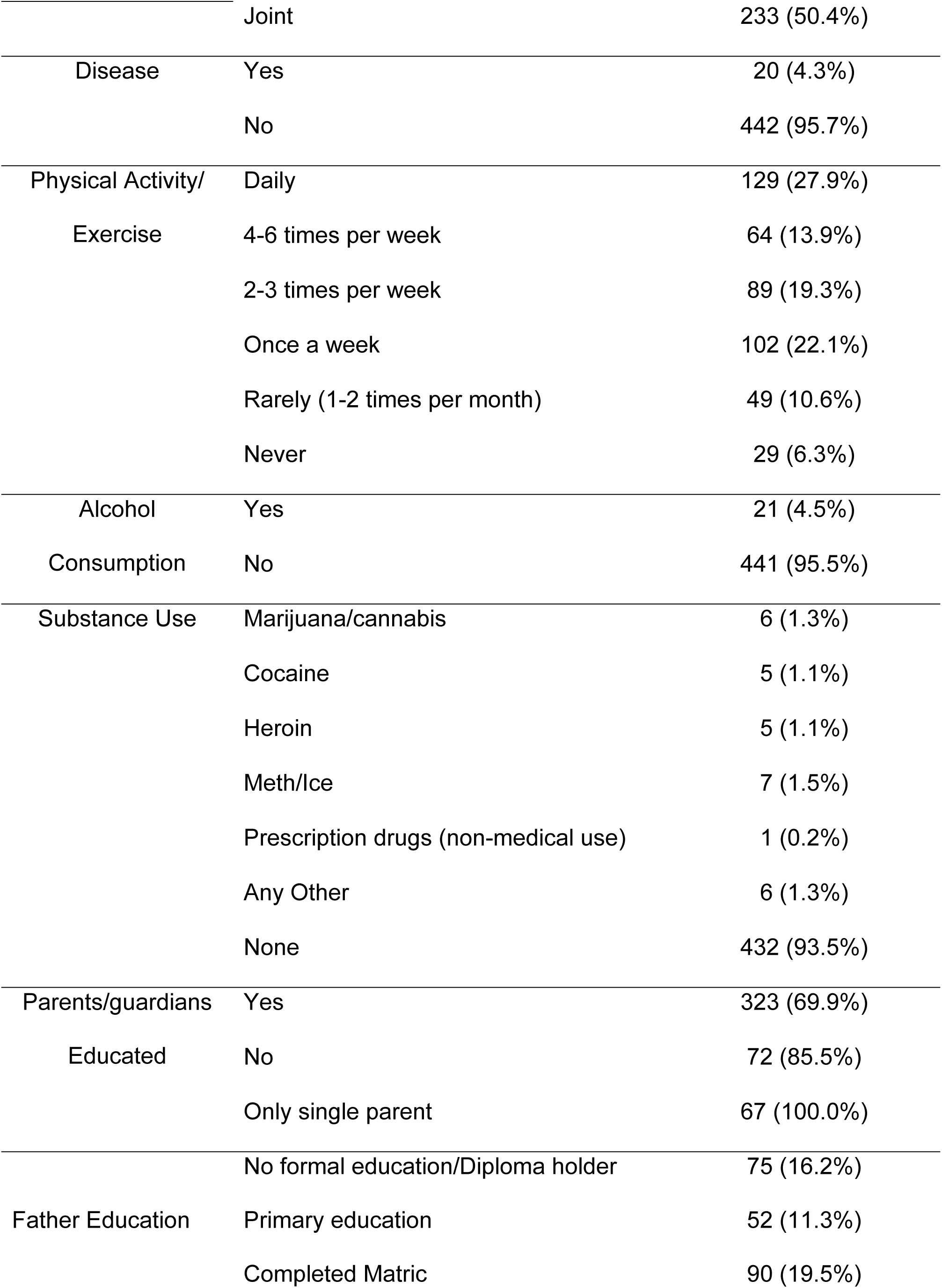
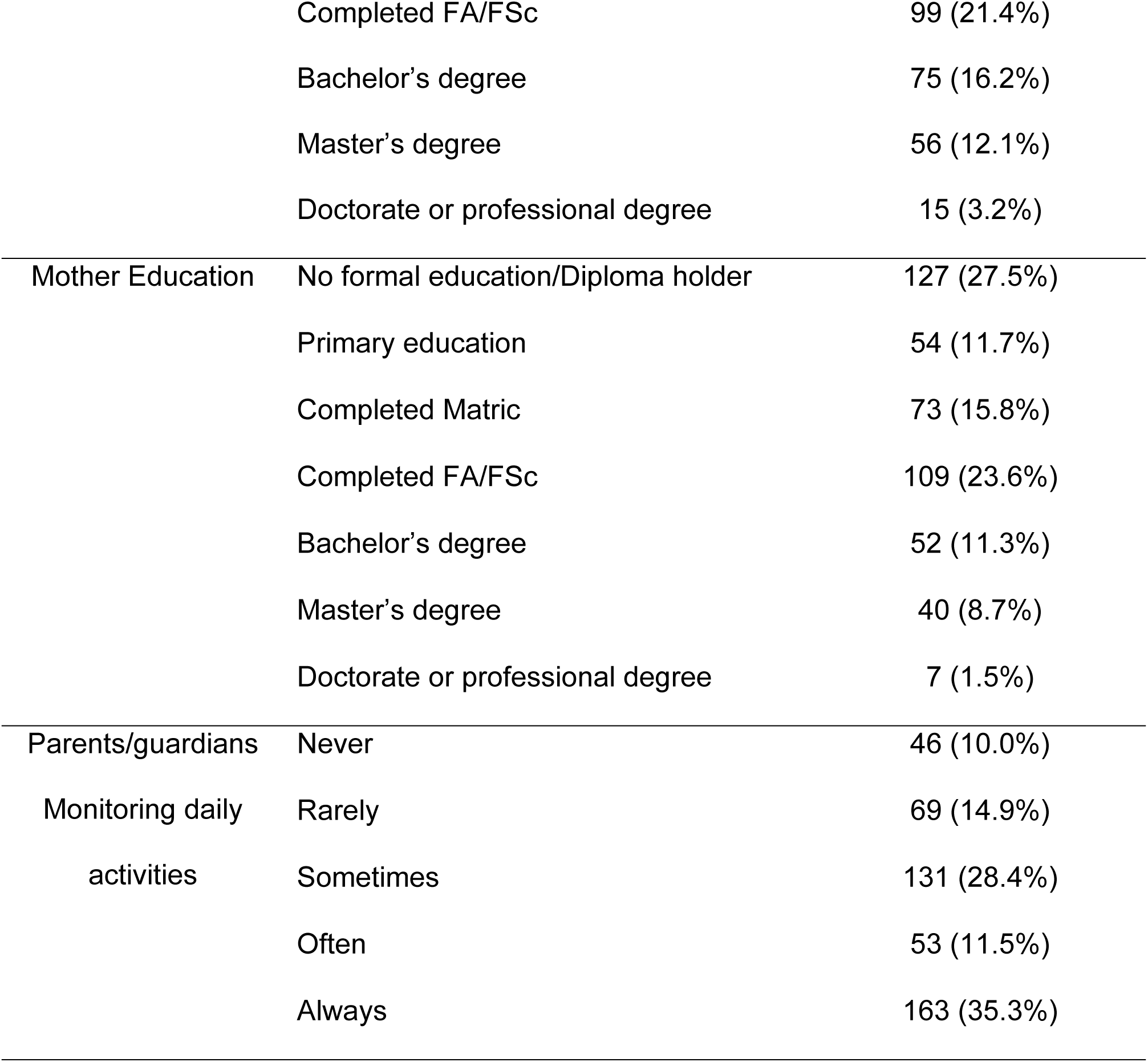
Demographics Characteristics of Respondents.

### Current Smoking Status among Youth

Table 2 presents the distribution of smoking status among the study population, with a total of 462 participants (N = 462). Most respondents were non-users/non-smokers (n = 248, 53.7%), indicating that more than half of the study population did not engage in either tobacco or E-cigarette use. Among those who reported current smoking behaviors, the most prevalent category was dual users, who simultaneously used both E-cigarettes and tobacco cigarettes (n = 109, 23.6%). Exclusive smoking behaviors were also evident, with current exclusive tobacco cigarette smokers comprising 20.6% (n = 95) of the sample. In contrast, only a small proportion of participants identified as current exclusive E-cigarette users (n= 10, 2.1%), indicating that sole reliance on e-cigarettes was far less common than either dual use or exclusive tobacco smoking (Table 2).

**Table 2.**
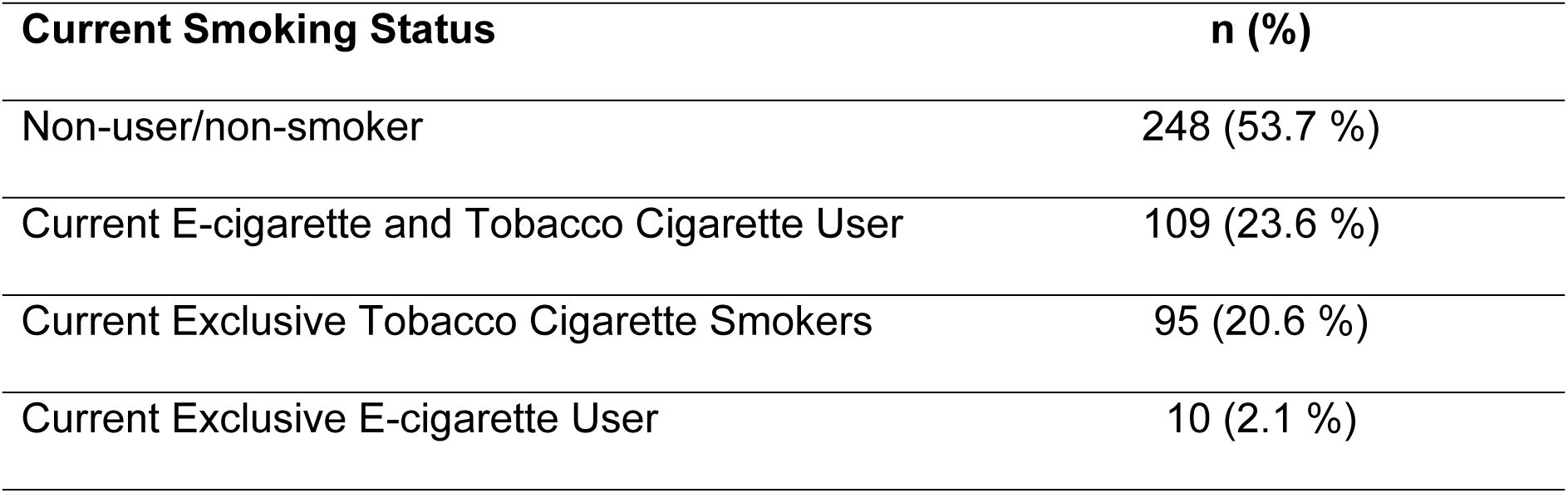
Current Smoking Status among Youth.

| **Current Smoking Status** | **n (%)** |
| --- | --- |
| Non-user/non-smoker | 248 (53.7 %) |
| Current E-cigarette and Tobacco Cigarette User | 109 (23.6 %) |
| Current Exclusive Tobacco Cigarette Smokers | 95 (20.6 %) |
| Current Exclusive E-cigarette User | 10 (2.1 %) |

### Patterns and Behaviors of Tobacco Consumption among Youth

Table 3 presents key findings on tobacco use, exposure, and cessation support among respondents. Of the 462 participants, 59.7% (n=276) tried smoking, while 40.3% (n=186) had never smoked. Smoking initiation was most common between ages 16– 18 (21%) and 22–24 (21.4%). In the past 30 days, 16.9% (n=78) smoked daily, while 15.6% (n=72) had not smoked. Flavored cigarette use was reported by 15% (n=70), with fruit flavors being the most preferred (9.1%). Besides cigarettes, the most used tobacco product was shisha (15.4%), followed by cigars (5.8%) and pipes (2.2%). Lower usage was reported for waterpipes (2.2%), hookahs (1.8%), bidis (1%), nicotine pouches (1.3%), and niswar (0.6%). Overall, 29% (n=134) had never tried any of these products, and 40.3% (n=186) had never smoked any form of tobacco. Regarding smoking cessation, 20.6% (n=95) received support from family or friends, 6.5% (n=30) sought help from a healthcare professional, and 3.5% (n=16) used formal cessation programs. School-based interventions (2.8%), digital resources (1.9%), and other sources (0.4%) were rarely utilized. Notably, 24% (n=111) received no cessation support. Media exposure to tobacco was high, with 41.1% (n=190) seeing it sometimes on TV/movies and 38.1% (n=176) on the internet/social media. Only 10% (n=46)reported exposure to tobacco advertisements at points of sale (Table 3).

**Table 3.**
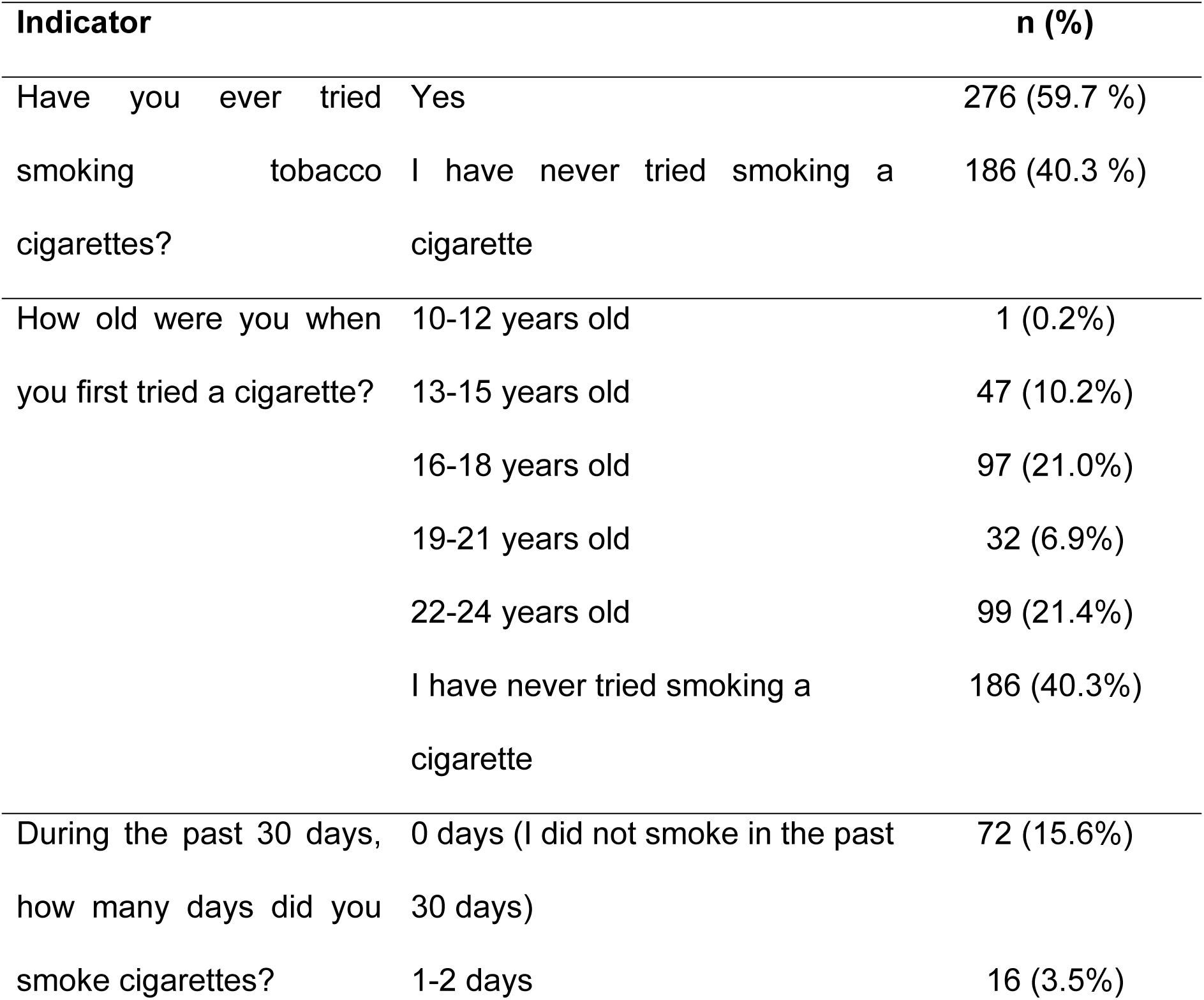
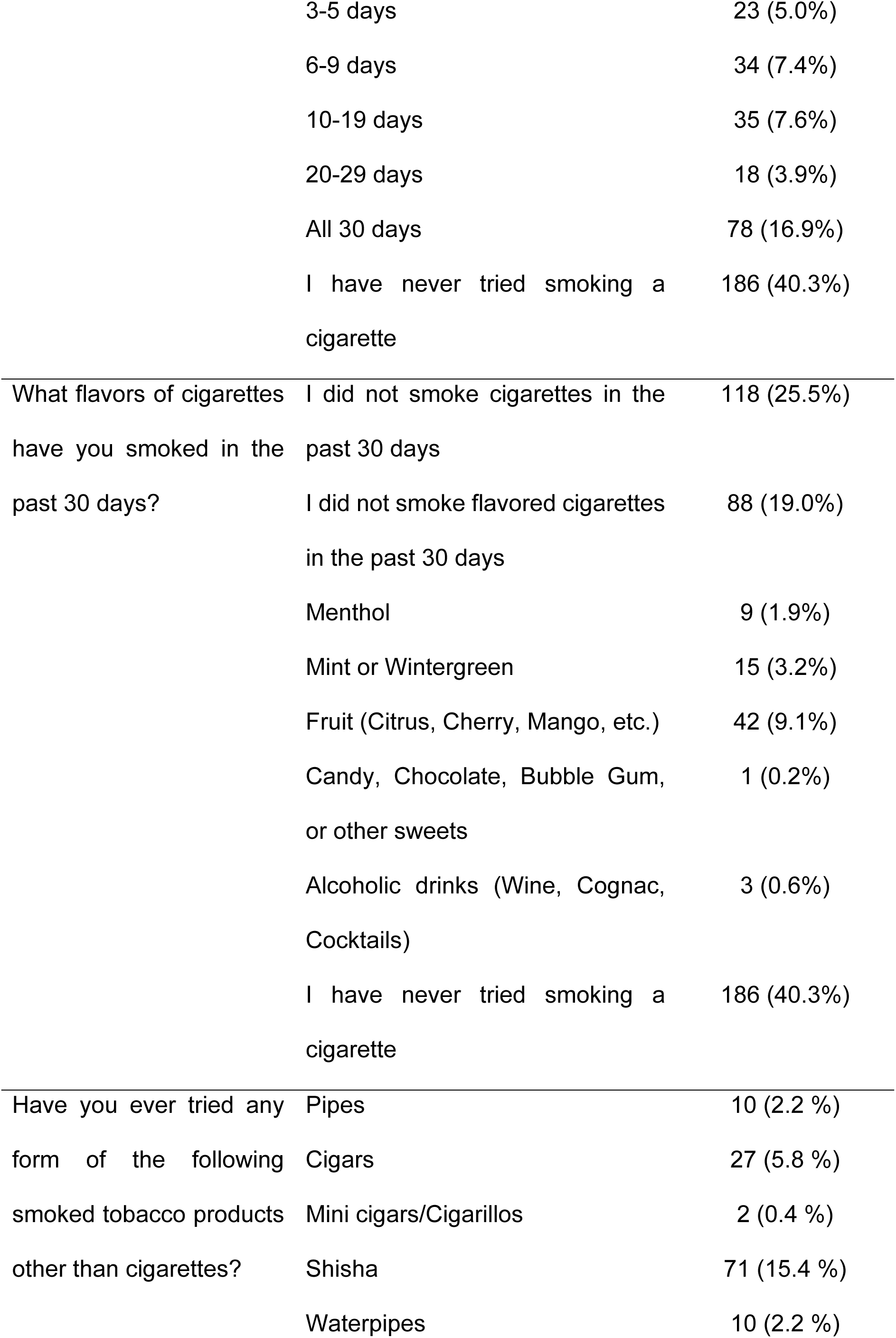
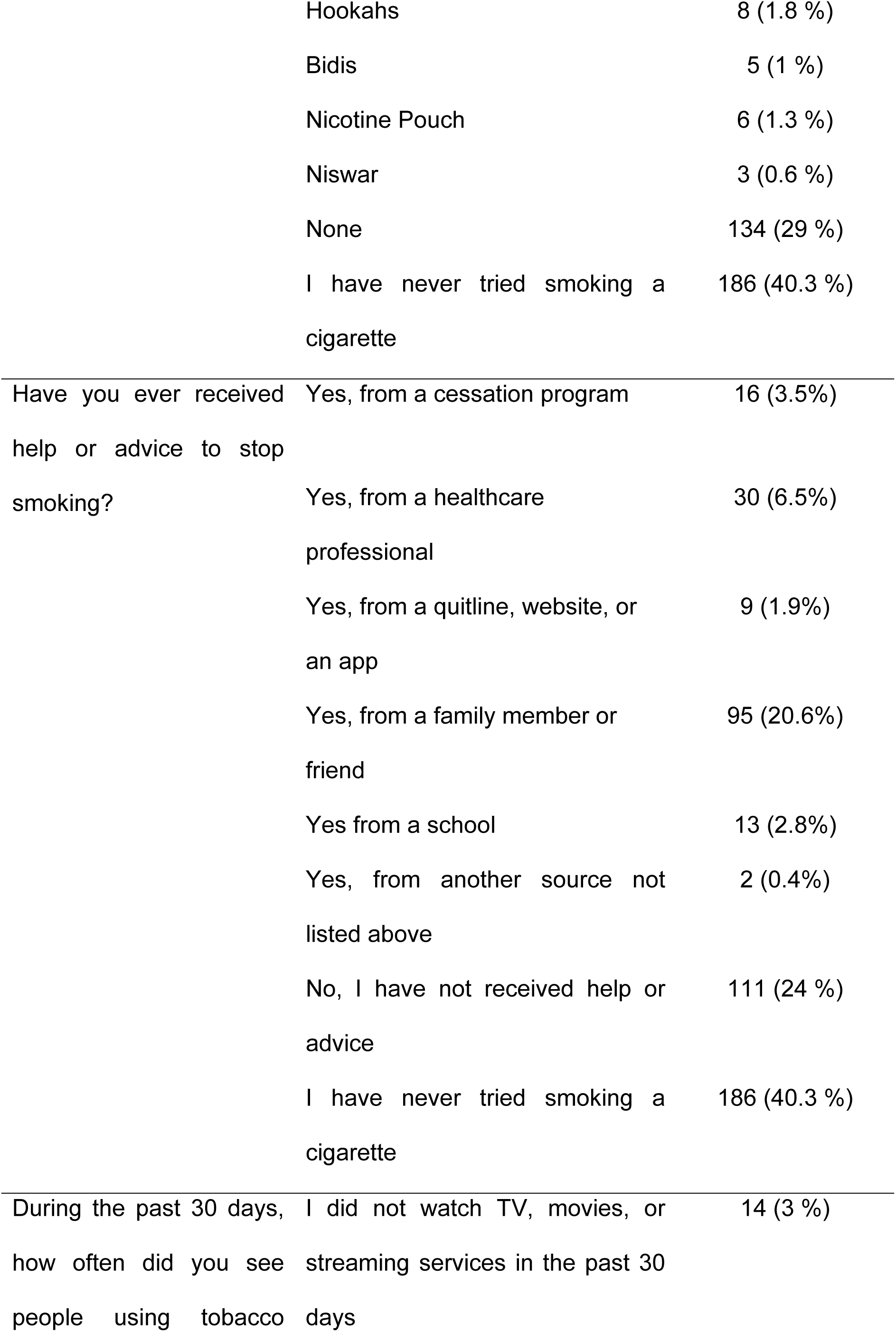
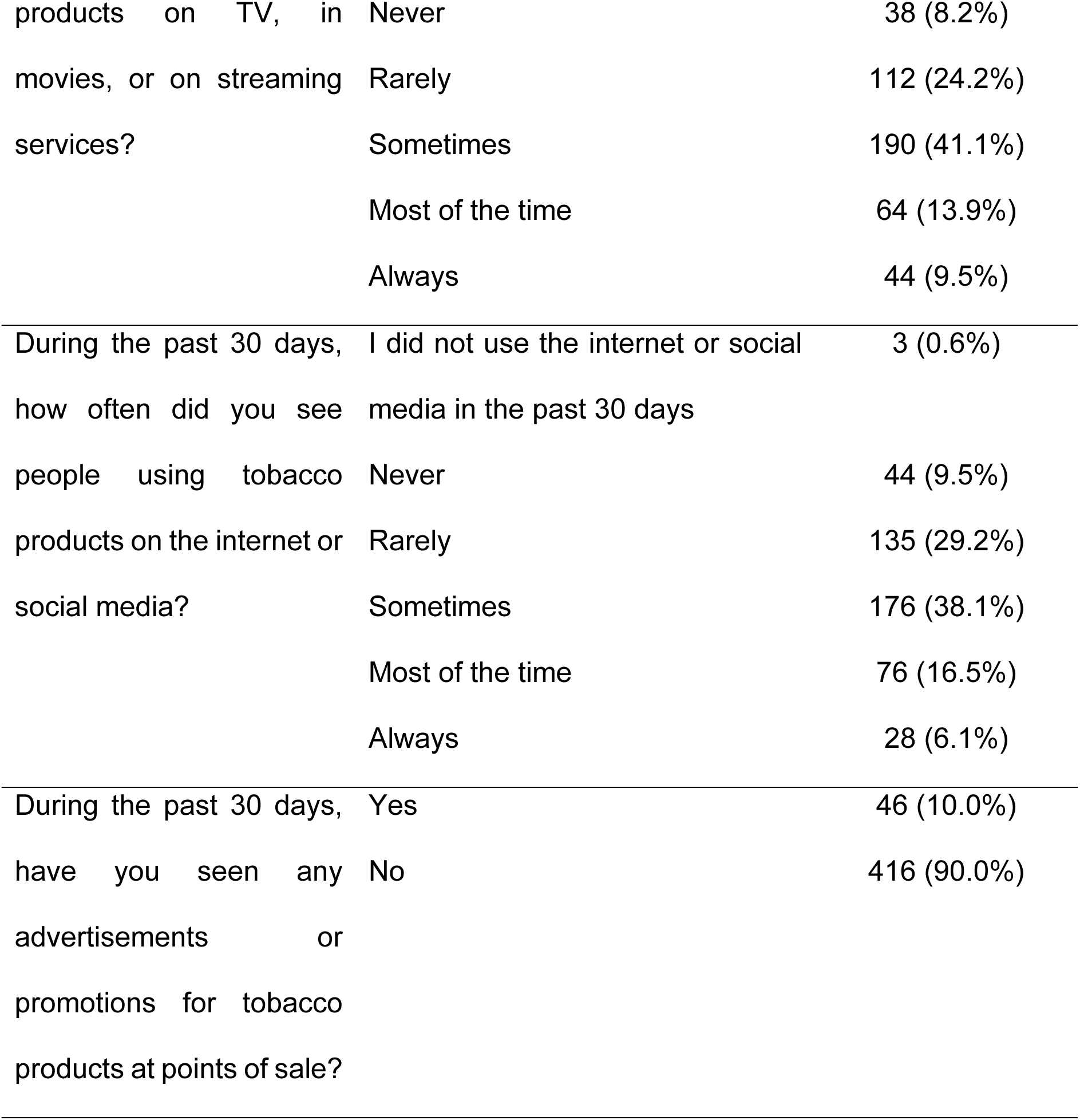
Patterns and Behaviors of Tobacco Consumption among Youth.

### Practice and Use Patterns of ENDS among Youth

Most of the respondents, 74.7% (n = 345) perceived E-cigarettes as easy or very easy to obtain for young people of their age, suggesting widespread availability despite regulatory controls. The primary reasons cited for initiating E-cigarette use included stress (31.4%, n = 145), depression (24 %, n = 111), with a smaller proportion attributing usage to peer pressure (9.1%, n = 42), social acceptability (9.1%, n = 42), recreational use (6.5%, n = 30), or smoking cessation attempts (10.8%, n = 50) while 7.3% (n = 34) of the respondents identified multiple combined factors as motivations for use, highlighting the complex interplay of psychological and social influences. Regarding social perceptions, 30.3% (n = 140) believed that young E-cigarette users have more friends, while 24.2% (n = 112) thought they have fewer friends, and nearly half (45.5%, n = 210) perceived no difference compared to non-users. Self-reported E-cigarette experimentation was noted among 25.8% (n = 119) of respondents, indicating a substantial level of exposure despite potential health risks. Perceived prevalence within social circles varied, with 24.0% (n = 111) believing no one among their peers uses E-cigarettes, while 50.8% (n = 235) estimated that between one and five out of every ten individuals in their grade, workplace, or neighborhood use E-cigarettes. Notably, 4.5% (n = 21) believed all ten peers use E-cigarettes, reflecting perceptions of normalization among certain groups (Table 5).

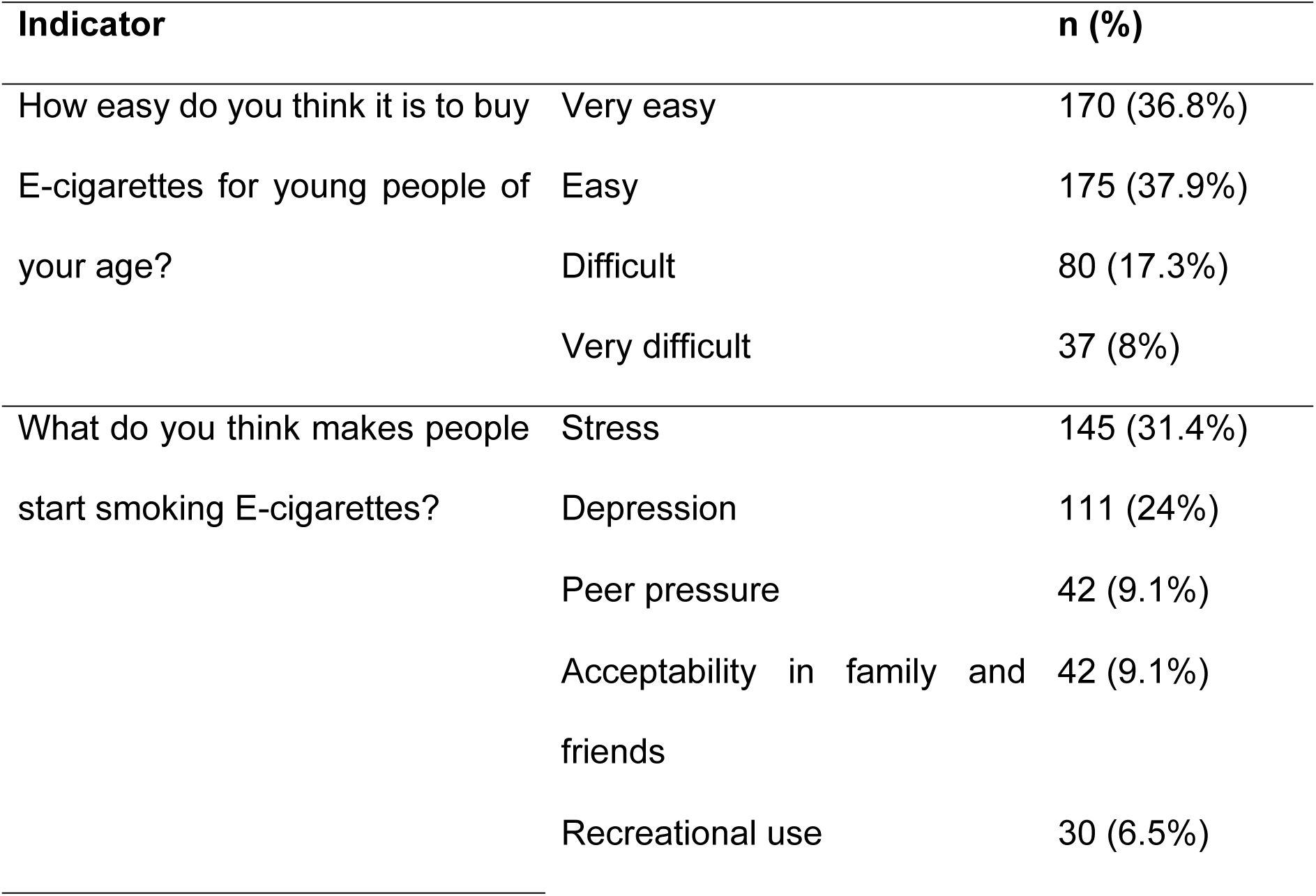
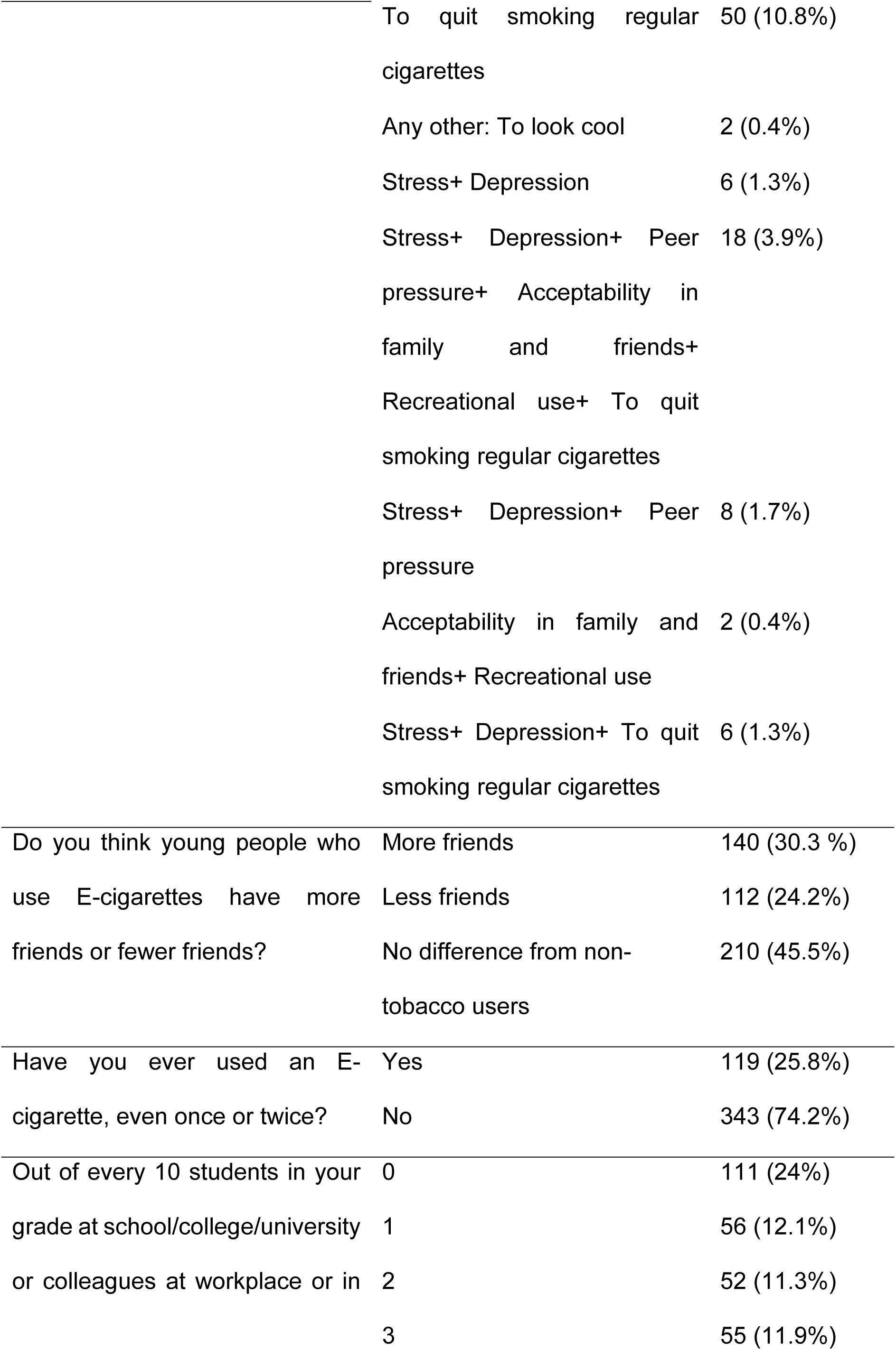
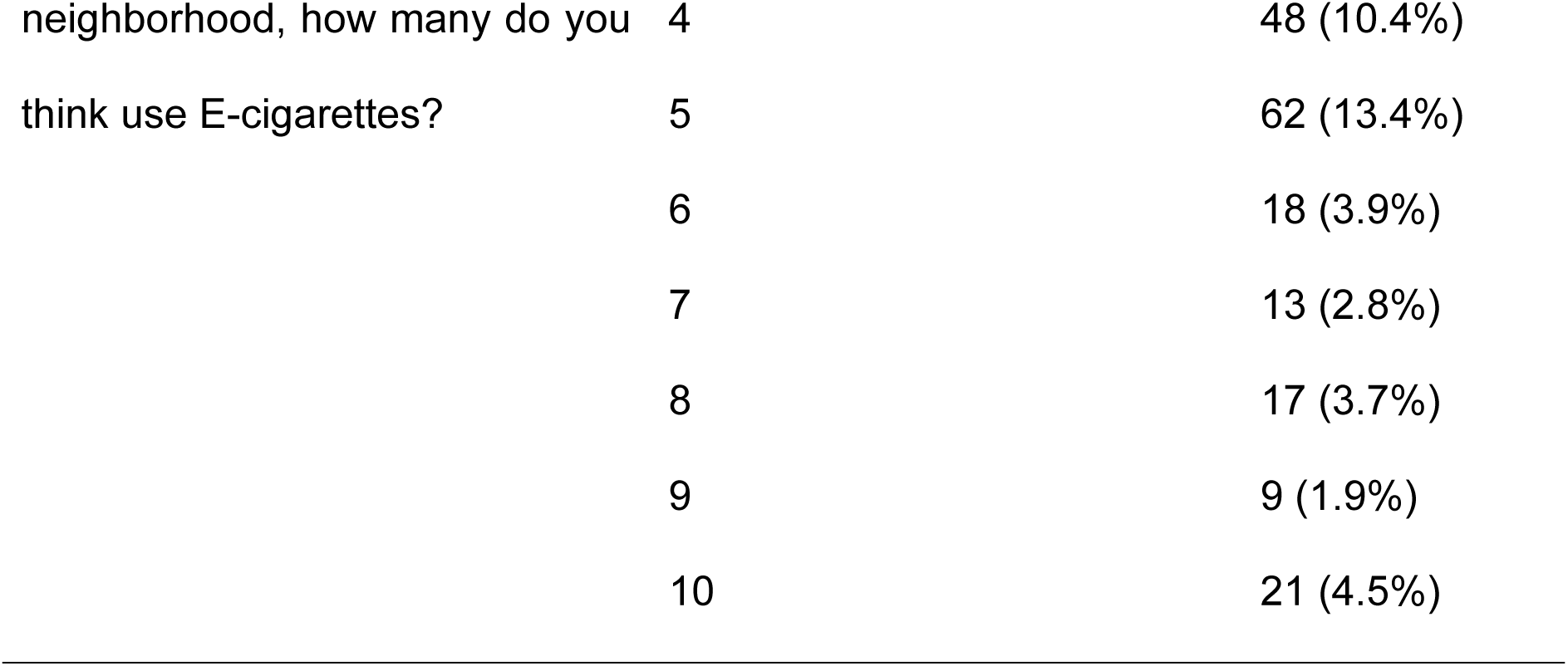

### Comparison of Demographic, Socioeconomic, and Behavioral Factors Among Nonsmokers, Dual Users, Exclusive Cigarette Users, and Exclusive ENDS Users

Chi-square analysis revealed significant associations between demographic, socioeconomic, and behavioral factors with smoking and ENDS use among youth. Younger individuals (15–17 years) were more likely to be nonsmokers or exclusive ENDS users, while older participants (23–25 years) had a higher prevalence of cigarette and dual use (p = 0.001). Males were significantly more likely to be dual, cigarette, or exclusive ENDS users, whereas females were more common among nonsmokers (p = 0.001). Exclusive ENDS use was concentrated in Islamabad, while cigarette use was more widespread in Punjab and Sindh (p = 0.001). Higher education levels (bachelor’s or master’s degrees) were more common among cigarette and dual users, whereas high school students had a higher proportion of nonsmokers (p = 0.001). Exclusive ENDS users were more likely from private and semi-government institutions, while public institutions had a higher proportion of nonsmokers (p = 0.029). Employment status (p = 0.001), higher household income (p = 0.001), and greater pocket money (p = 0.001) were associated with increased ENDS and cigarette use. Exclusive ENDS users were more likely to live with parents, whereas dual and cigarette users commonly resided in shared accommodations or hostels (p = 0.011). Alcohol (p = 0.045) and substance use (p= 0.001) were significantly higher among tobacco and ENDS users. Parental monitoring was a protective factor, with nonsmokers reporting the highest supervision levels (p = 0.036). Birth order, family structure, health conditions, and physical activity showed no significant associations. These findings highlight that age, gender, education, income, employment, lifestyle behaviors, and parental supervision strongly influence smoking and ENDS use among youth (Table 6).

**Table 6.**
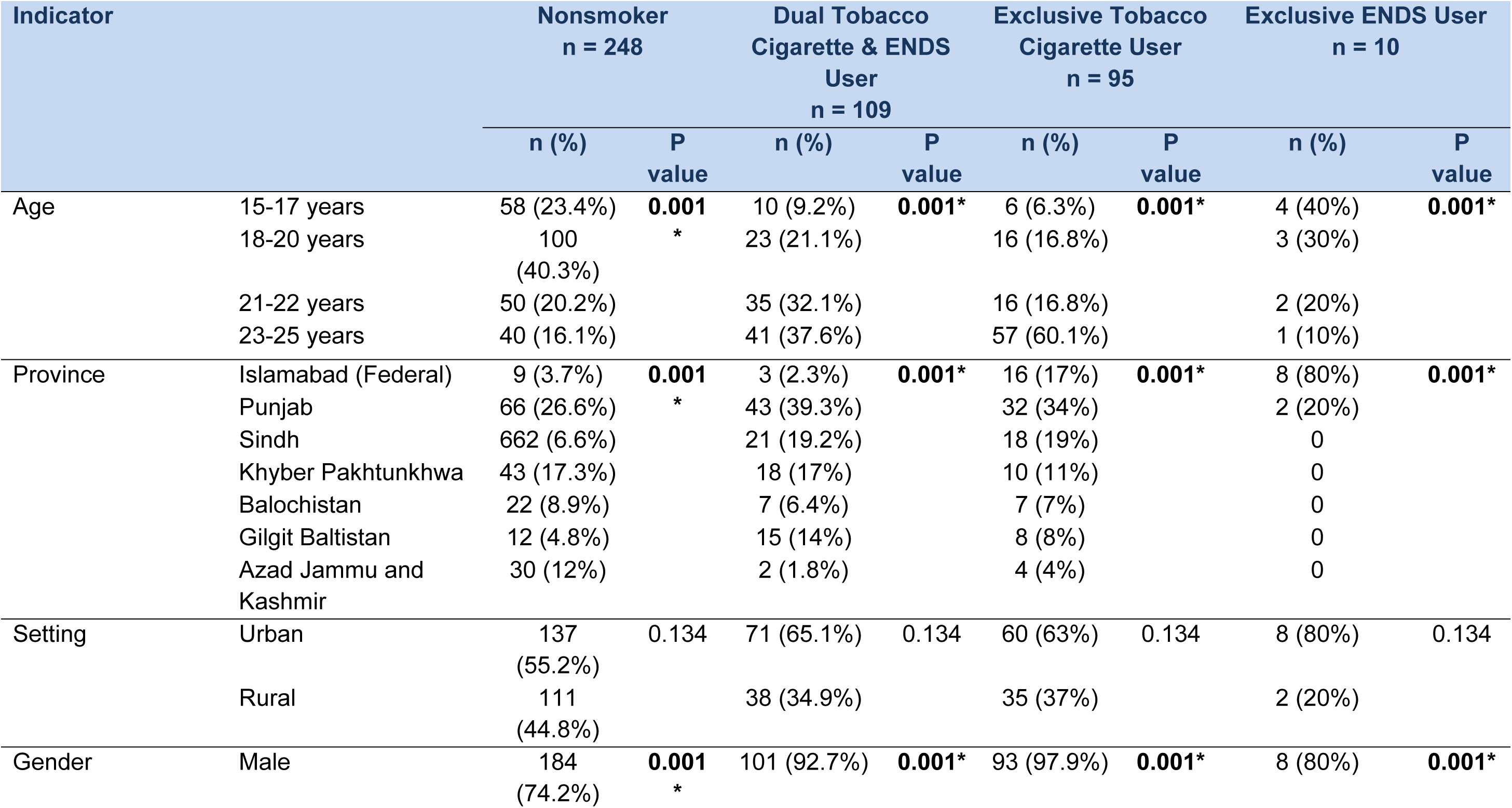
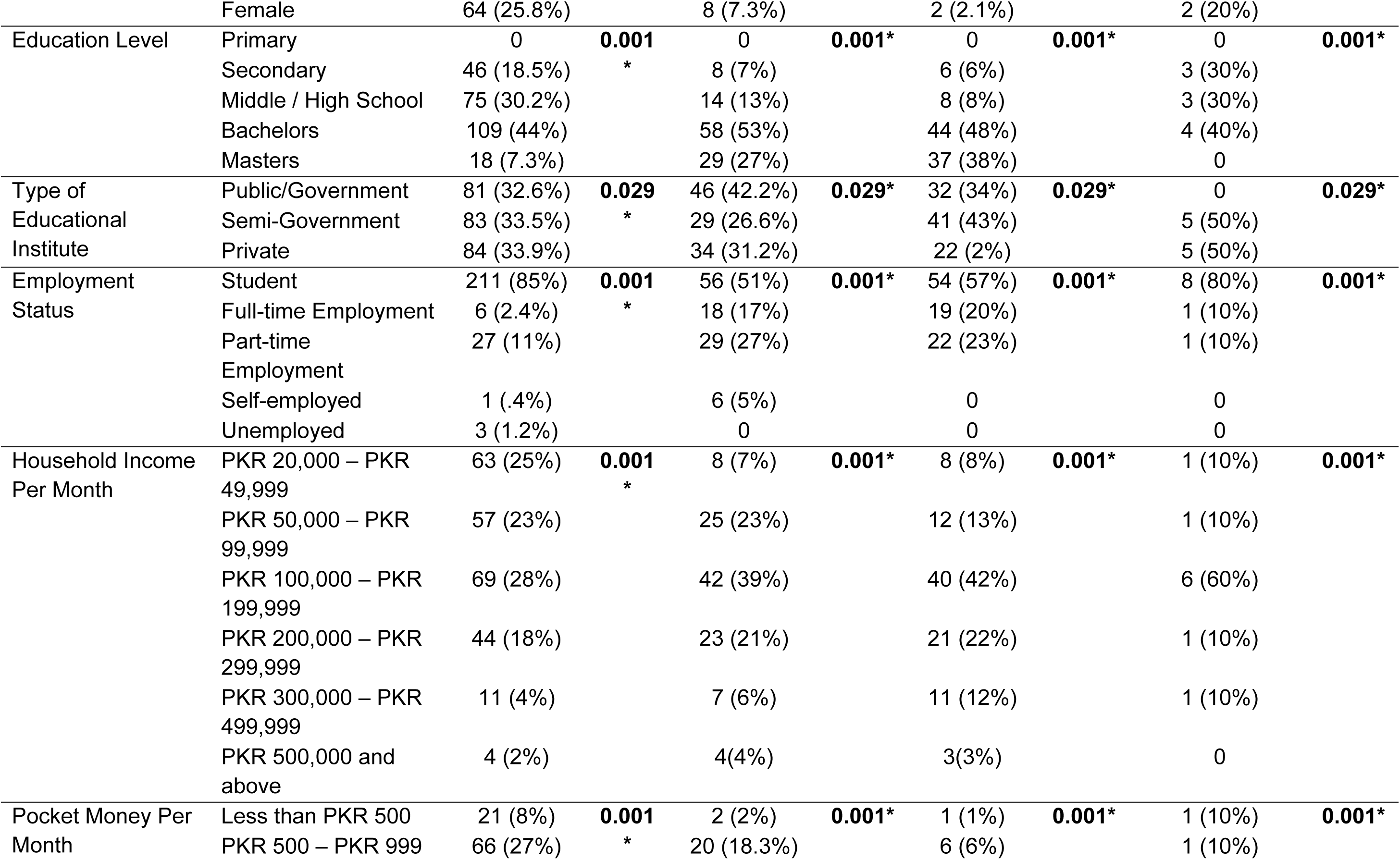
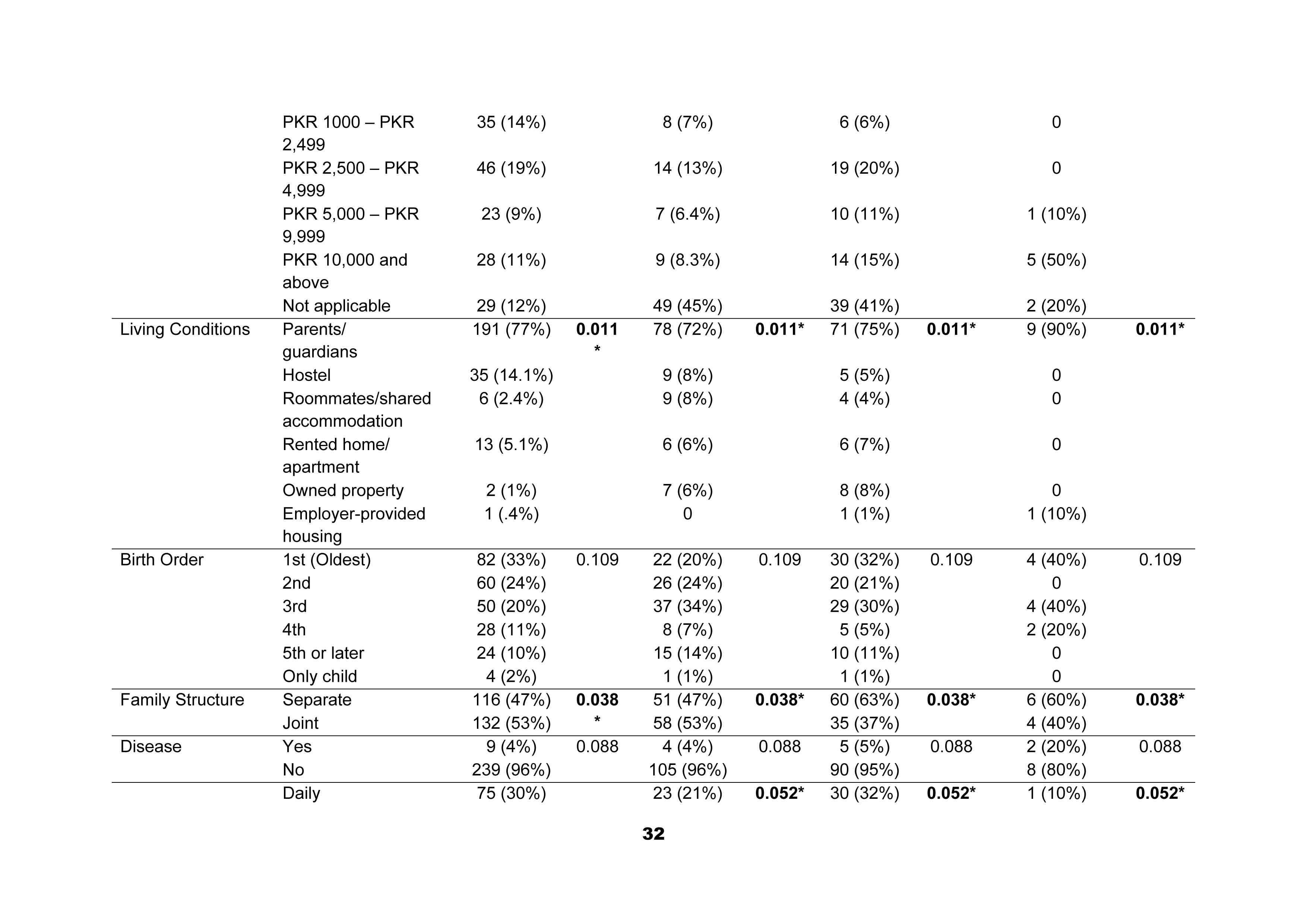
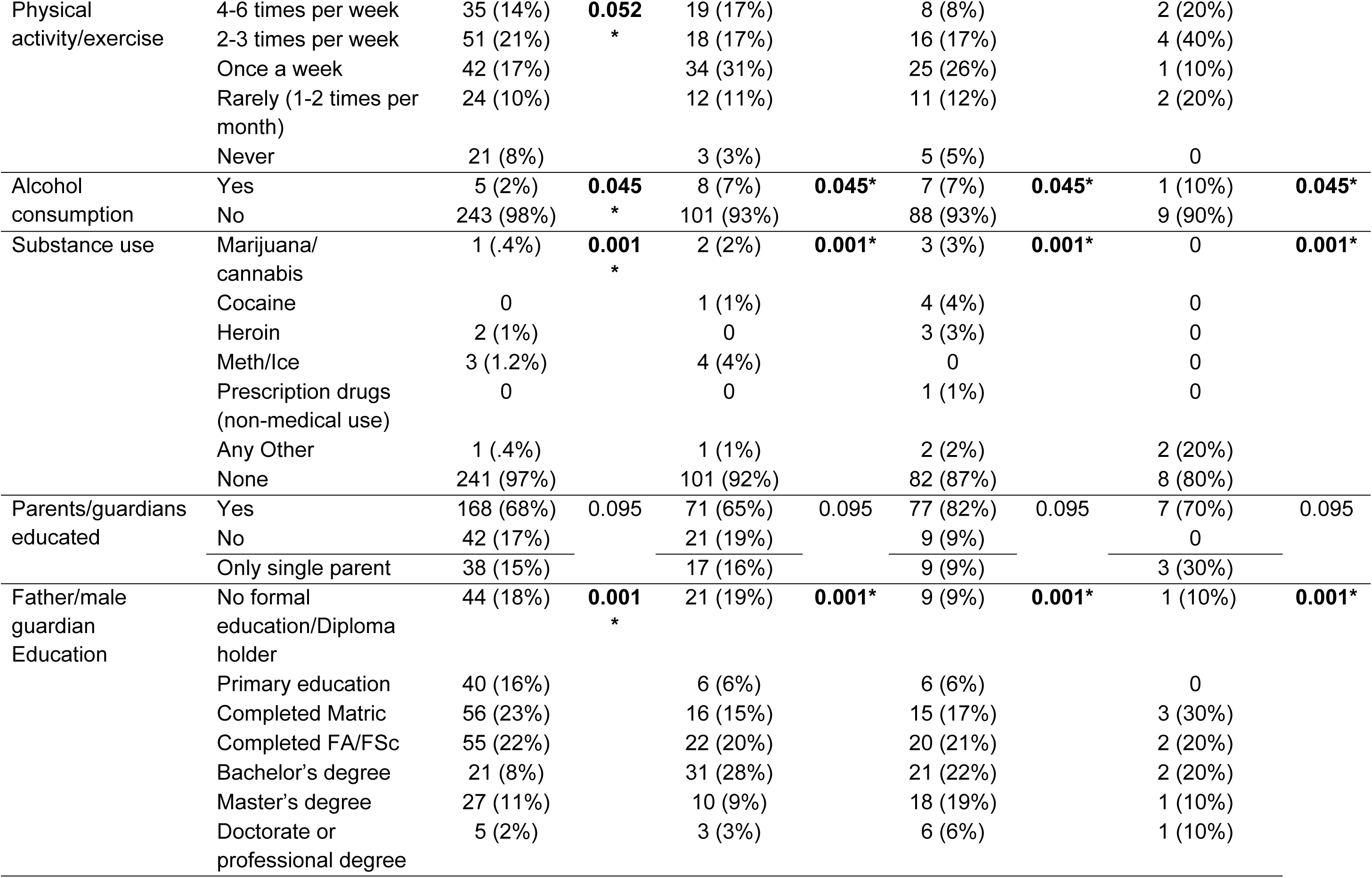
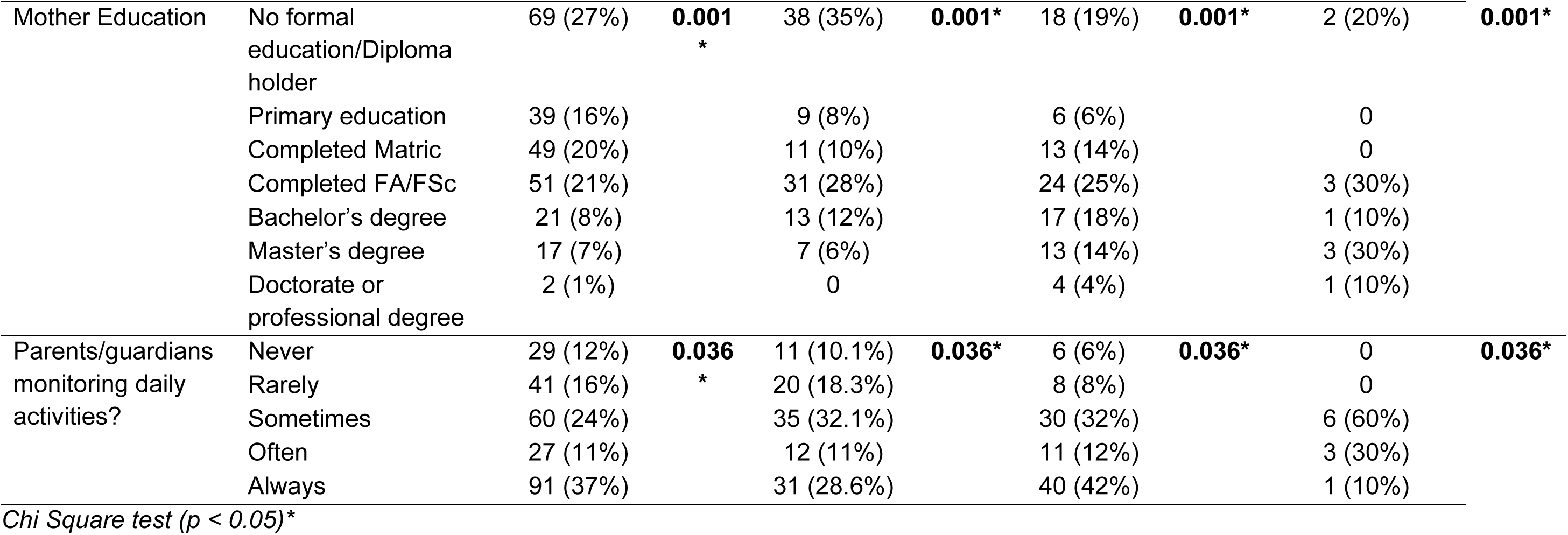
Comparison of Demographic, Socioeconomic, and Behavioral Factors Among Nonsmokers, Dual Users, Exclusive Cigarette Users, and Exclusive ENDS Users.

### Relationship Between Knowledge, Attitude, and Smoking Behavior

Spearman’s correlation analysis was conducted to examine the relationships between knowledge, attitude, and smoking status regarding ENDS. The results indicate a weak but statistically significant positive correlation between knowledge and attitude (r⍰ = 0.300, p < 0.001), suggesting that individuals with greater knowledge about ENDS tend to have more favorable attitudes toward them. Additionally, a weak negative correlation was found between knowledge and smoking status (r⍰ = -0.137, p = 0.003), indicating that higher knowledge levels are slightly associated with a lower likelihood of smoking. However, no significant correlation was observed between attitude and smoking status (r⍰ = -0.006, p = 0.895), implying that an individual’s attitude toward ENDS does not strongly predict their smoking behavior. These findings suggest that while knowledge plays a role in shaping attitudes, its impact on smoking behavior is limited, highlighting the need for comprehensive educational and behavioral interventions to address ENDS use effectively (Table 7).

**Table 7.**
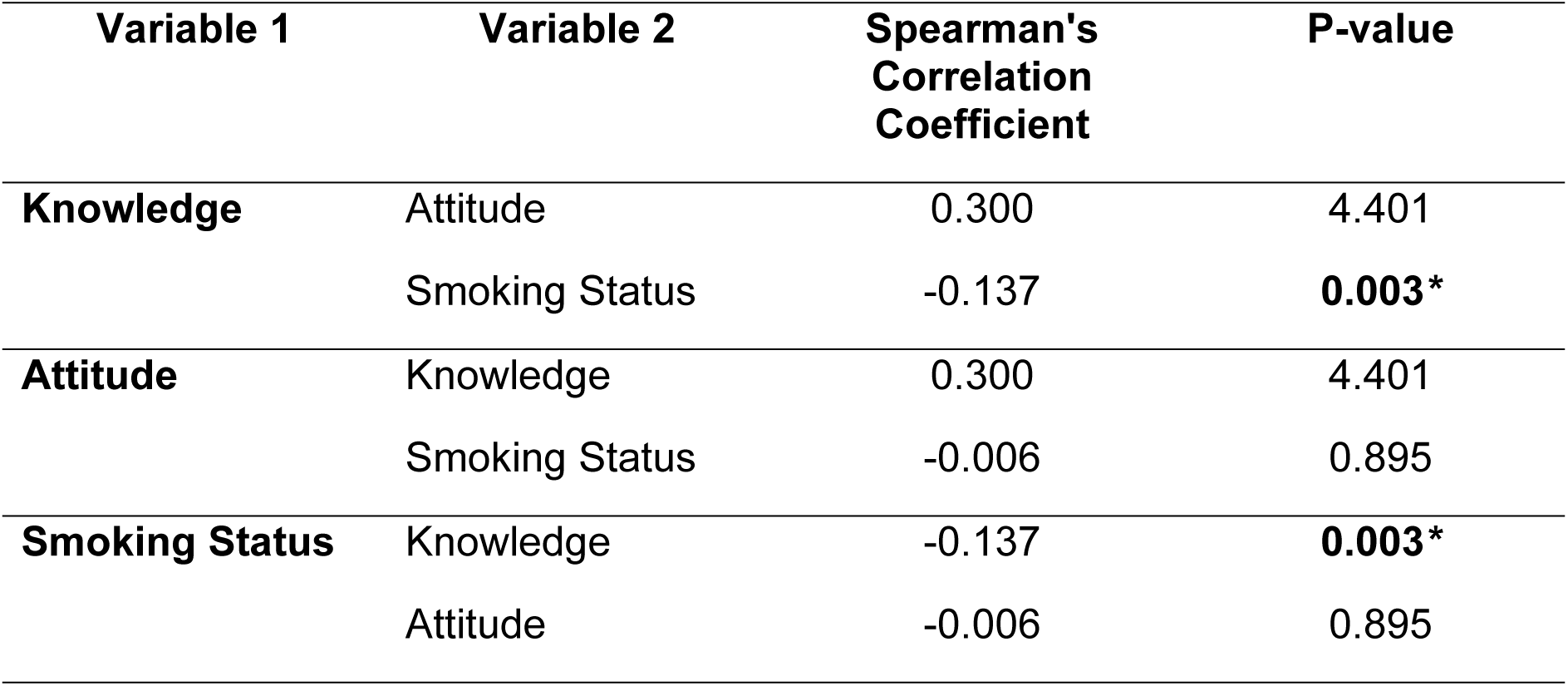
Relationship Between Knowledge, Attitude, and Smoking Behavior.

| Variable 1 | Variable 2 | Spearman's Correlation Coefficient | P-value |
| --- | --- | --- | --- |
| **Knowledge** | Attitude | 0.300 | 4.401 |
|  | Smoking Status | -0.137 | **0.003\*** |
| **Attitude** | Knowledge | 0.300 | 4.401 |
|  | Smoking Status | -0.006 | 0.895 |
| **Smoking Status** | Knowledge | -0.137 | **0.003\*** |
|  | Attitude | -0.006 | 0.895 |

### Impact of Demographic and Behavioral Factors on Youth Knowledge Regarding ENDS

The multivariate analysis identified key predictors, risk factors, and protective factors influencing youth knowledge and attitudes toward ENDS. Females exhibited higher knowledge levels than males (AOR = 5.22, p = 0.009), and urban residency was also linked to greater awareness (AOR = 1.44, p = 0.0125). Employment status emerged as a protective factor, with working youth having higher knowledge levels (AOR = 0.43, p = 0.0146). However, substance use was associated with lower knowledge (AOR = 1.57, p = 0.0565), while regional differences lost significance after adjustment. Factors such as education level, parental education, income, marital status, physical activity, alcohol consumption, and smoking status had no significant impact on knowledge.

Regarding attitudes, older youth were more likely to hold favorable perceptions of ENDS (AOR = 1.63, p = 0.0401), and urban residents exhibited higher odds of positive attitudes (AOR = 1.34, p = 0.0328). However, regional differences (AOR= 0.44, p = 0.0071), employment (AOR = 0.40, p = 0.0096), and physical activity (AOR = 0.66, p = 0.0004) served as protective factors against favorable attitudes toward ENDS. Gender, education level, household income, parental education, substance use, smoking status, and alcohol consumption did not significantly influence attitudes (Table 8).

**Table 8.** Impact of Demographic and Behavioral Factors on Youth Knowledge Regarding ENDS.

| Variable | Knowledge |  |  |  |  |  | Attitude |  |  |  |  |  |
| --- | --- | --- | --- | --- | --- | --- | --- | --- | --- | --- | --- | --- |
|  | Bivariate |  |  | Multiple |  |  | Bivariate |  |  | Multiple |  |  |
|  | OR | 95% CI | P-value | AOR | 95% CI | P-value | OR | 95% CI | P-value | AOR | 95% CI | P-value |
| Age | 0.8 | (0.55, 1.15) | 0.466 | 0.8 | (0.5, 1.29) | 0.362 | 1.58 | (1.09, 2.29) | 0.052 | 1.63 | (1.02, 2.59) | 0.040 |
| Gender | 3.34 | (1.16, 9.61) | **0.035\*** | 5.22 | (1.51, 18.0) | **0.009\*** | 0.75 | (0.3, 1.91) | 0.716 | 0.87 | (0.29, 2.66) | 0.812 |
| Province | 1.36 | (1.13, 1.65) | **0.001\*** | 0.68 | (0.36, 1.26) | 0.216 | 0.83 | (0.69, 0.99) | 0.052 | 0.44 | (0.24, 0.8) | 0.007 |
| Setting | 1.19 | (1.09, 1.29) | **0.002\*** | 1.44 | (1.08, 1.92) | **0.012\*** | 0.94 | (0.87, 1.02) | 0.237 | 1.34 | (1.02, 1.76) | 0.032 |
| Marital Status | 0.49 | (0.04, 5.49) | 0.992 | 0.15 | (0.01, 2.35) | 0.175 | 0.0 | (0.0, inf) | 0.225 | 0.04 | (0.0, 22.27) | 0.319 |
| Education | 0.95 | (0.53, 1.71) | 0.360 | 1.34 | (0.61, 2.96) | 0.467 | 1.47 | (0.81, 2.68) | 0.501 | 1.21 | (0.55, 2.66) | 0.634 |
| Employment | 0.69 | (0.5, 0.95) | 0.154 | 0.43 | (0.22, 0.85) | **0.014\*** | 0.63 | (0.46, 0.87) | 0.030 | 0.4 | (0.2, 0.8) | 0.009 |
| House Income | 0.76 | (0.58, 0.99) | 0.224 | 0.81 | (0.57, 1.13) | 0.210 | 0.97 | (0.75, 1.25) | 0.015 | 1.08 | (0.77, 1.5) | 0.656 |
| Own Income | 1.09 | (0.96, 1.24) | 0.596 | 0.8 | (0.61, 1.05) | 0.103 | 1.13 | (1.0, 1.28) | 0.097 | 0.84 | (0.64, 1.1) | 0.196 |
| Physical Activity | 0.97 | (0.8, 1.16) | 0.752 | 0.9 | (0.72, 1.13) | 0.368 | 0.72 | (0.59, 0.87) | 0.002 | 0.66 | (0.52, 0.83) | 0.000 |
| Alcohol Use | 0.49 | (0.09, 2.77) | 0.687 | 0.16 | (0.02, 1.65) | 0.123 | 2.16 | (0.39, 12.09) | 0.631 | 1.3 | (0.16, 10.73) | 0.805 |
| Substance Use | 1.34 | (0.98, 1.84) | 0.187 | 1.57 | (0.99, 2.49) | **0.056\*** | 1.33 | (0.98, 1.8) | 0.183 | 1.27 | (0.89, 1.81) | 0.192 |
| Smoking Status | 0.96 | (0.74, 1.23) | 0.961 | 1.01 | (0.73, 1.39) | 0.959 | 0.96 | (0.74, 1.24) | 0.465 | 0.96 | (0.7, 1.32) | 0.794 |
| Parents Education | 1.47 | (0.96, 2.27) | 0.182 | 1.42 | (0.69, 2.9) | 0.341 | 1.07 | (0.71, 1.63) | 0.214 | 0.97 | (0.5, 1.88) | 0.918 |
| Father Education | 0.82 | (0.69, 0.98) | **0.001\*** | 0.88 | (0.66, 1.17) | 0.375 | 0.94 | (0.79, 1.12) | 0.030 | 0.92 | (0.7, 1.21) | 0.561 |
| Mother Education | 0.83 | (0.69, 0.99) | **0.046\*** | 1.14 | (0.78, 1.69) | 0.495 | 0.93 | (0.78, 1.1) | 0.315 | 0.97 | (0.68, 1.37) | 0.852 |
| Family Structure | 0.98 | (0.55, 1.74) | 1.0 | 0.95 | (0.47, 1.93) | 0.895 | 0.9 | (0.51, 1.61) | 0.842 | 0.84 | (0.41, 1.7) | 0.627 |

*Multivariate logistic regression analysis ( $p < 0.05$ )\**

## Discussion

Understanding the factors that influence ENDS and cigarette use is essential for developing effective prevention and intervention strategies. This study highlights significant knowledge gaps and common misconceptions about ENDS, indicating that awareness alone cannot readily decrease tobacco use. Attitudes toward smoking and ENDS are directly related to positive perceptions increasing the likelihood of use, reinforcing the impact of social acceptance and marketing strategies. The findings of this study highlight significant knowledge gaps among youth regarding the health risks, nicotine content, and regulatory policies of ENDS, contributing to increased exposure to their use. A considerable proportion of participants believe that ENDS do not contain nicotine, or they are less harmful than traditional cigarettes. Similarly, Willett et al. (2019) and Mantey et al. (2019), reported that marketing strategies portraying ENDS as "clean" alternatives to cigarettes have led to extensive misinformation among adolescents ^[4, 11]^. The present study further revealed that urban youth had higher knowledge levels as compared to their rural counterparts. Similar trend was observed by Leventhal et al. (2019) ^[5]^. However, despite this increased awareness, urban youth also showed higher rates of ENDS use, emphasizing findings from Kowitt et al. (2019) that knowledge alone does not effectively prevent tobacco use when thwarted by strong social acceptability and accessibility ^[12]^.

The present study demonstrated marked gender disparities in knowledge, with females exhibiting greater awareness of ENDS risks than males. This aligns with research by Alves et al. (2022), which found that females are more likely to engage with health education initiatives and exhibit greater risk aversion ^[13]^. However, despite higher awareness levels, female smoking prevalence remained significantly lower than that of males, suggesting that cultural norms may serve as a stronger protective factor than knowledge alone. In contrary, the smoking rates were much higher in males, with lower levels of knowledge, which is consistent with the findings of Jha et al. (2020) that peer reinforcement and sociocultural factors have a preponderant role in smoking initiation among young men ^[14]^. Likewise, the current study found that the employed group knew more about ENDS, possibly because of workplace health awareness programs and social exposure to tobacco users. However, higher levels of knowledge did not lead to reduced prevalence of smoking, supporting earlier findings by Choi and Forster (2013), which found that greater financial independence is associated with higher tobacco use due to less parental oversight and increased purchasing capacity ^[15]^.

Demographic and behavioral characteristics had a strong influence on attitudes towards ENDS, with urban youth exhibiting more positive perceptions compared to their rural counterparts. These findings are in line with the research by Cullen etal. (2018), which showed that urban youth are more exposed to ENDS advertisements, peer influence, and easier retail accessibility, all of which contribute to increased acceptance of ENDS use ^[2]^. Urban youth, despite better knowledge level, also endorsed the perception that ENDS are fashionable compared to their rural counterparts, supporting findings from Leventhal et al. (2019) that social desirability is vital for the continuity of tobacco consumption ^[5]^. Similar to previous research by Wang et al. (2020), the present study found that older youth demonstrated more positive attitudes toward ENDS compared to younger participants ^[16]^. This might be due to increased exposure to environments where smoking is socially acceptable as the age increases, making it harder to counteract pro-tobacco attitudes.

Furthermore, the present study found noticeable gender differences in attitudes towards ENDS, with males significantly more likely to have positive perceptions of ENDS. These findings are consistent with research from Islam et al. (2020), which highlighted that sociocultural norms deter women from smoking leading to more negative attitudes toward tobacco among women ^[7]^. The current study found parental supervision also as an important determinant of attitudes, where increased parental supervision was associated with more negative attitudes toward smoking and use of ENDS. These findings were similar to trends reported by a study by Alves et al. (2021), providing evidence that strong parental involvement is an important protective factor for adolescent experimentation with tobacco ^[13]^.

Demographic, socioeconomic, and behavioral factors were significantly associated with smoking behavior. Sex was strongly and significantly associated with smoking status, with males smoking significantly more than females, similar to findings reported by Jha et al. (2020) ^[14]^. Financial independence was another major predictor of smoking behavior, reporting employed youth smoke more than non-working youth, which has also been reported by Leventhal et al. (2019) ^[5]^. Household income also affected tobacco use, with youth having higher-income households were found more likely to be ENDS users, and youth in lower-income households were more likely to smoke cigarettes. These findings are in line with the results reported by Mantey et al. (2019) and Willemsen et al. (2019) which found that ENDS are predominantly marketed as premium lifestyle products and are more often affordable for more affluent individuals while regular cigarette use still predominates the middle- and lower-income groups ^[11, 17]^.

The present study identified behavioral risk factors including alcohol use and substance use as strong predictors of smoking and ENDS use. The relationship between use of tobacco and alcohol is well-established showing that these substances commonly co-occur due to their reinforcing effects in social contexts (Jackson et al. (2021) ^[18]^. Similarly, smoking and dual use populations also reported higher use of substances such as marijuana, cocaine, and methamphetamine later in life, supporting the findings of Wills et al., (2016) which showed that ENDS and cigarettes tend to function as gateway substances to polysubstance use ^[19]^. These data indicate that smoking prevention interventions should be delivered as part of a broader substance abuse effort targeting several risk behaviors concurrently rather than tobacco use alone. One of the main findings of this study was that ENDS users were less likely to dual use with cigarettes than traditional smokers, which indicates that ENDS may act as an auxiliary product rather than an additional product. Research by Wills et al. (2016) suggest that while ENDS are indeed perceived as a potential gateway pathway for non-smokers, among current smokers, ENDS use is associated with more potential harm reduction than with increasing tobacco dependence ^[19]^. This contradicts the traditional narrative that all users of ENDS are at increased risk of smoking cigarettes and emphasizes the need to differentiate between users who have never smoked and those who are smoking cessation aids.

The current study showed that parental monitoring emerged as one of the most significant preventive factors of smoking, as nonsmokers reported higher levels of parental monitoring. These findings are consistent with the study by Wang et al. (2020), which indicated strong parental oversight delayed smoking initiation and more so, led to less susceptibility to peer pressure ^[16]^. The present study also confirmed being student as a protective factor against tobacco use supporting findings from Arrazola et al. (2015) that structured educational environments are discouraging smoking by advocating anti-tobacco norms ^[20]^. However, physical activity was not strongly related to smoking, suggesting that while exercise is healthy, it does not deter tobacco use unless targeted anti-smoking measures are also taken. This aligns with the research conducted by Audrain-McGovern et al. (2013) which found an inverse association between smoking and physical activity i.e. people engaging in physical activity are less likely to be smokers, the effect is minimal unless supported by behavioral reinforcement strategies ^[21]^.

The present study revealed that higher knowledge levels were related to more negative attitudes toward ENDS, but these attitudes did not necessarily relate to lower smoking prevalence. These results are consistent with the earlier work done by Kowitt et al. (2019) and Kwon et al. (2020) ^[12, 22]^. A weak negative correlation was found in this study between knowledge and smoking behavior which might due to external factors, such as peer pressure, social acceptability of smoking (in the case of tobacco), and stress-related motivations to engage smoking behavior in non-daily users. Leventhal et al. (2019) found that although adolescents with knowledge of smoking’s dangers may express negative attitudes toward smoking, possible use of tobacco can occur under social and environmental reinforcement. This highlights the need for the inclusion of behavioral interventions along with knowledge-based programs to effectively reduce youth smoking ^[23]^.

This present study reported that young people who perceive ENDS as less harmful were more likely to report using them even if they were aware of their possible harm. These results are in line with the findings reported by Willett et al. (2019) ^[4]^. The results of the current study are also in line with findings from Islam et al. (2020), which revealed that marketing, peer influence, and perceived lifestyle benefits of ENDS informed the emergence of positive attitudes toward smoking ^[7]^. These findings emphasize the importance of increased awareness and change of social norms and tackling the positive perceptions associated with smoking through counter-marketing campaigns, peer-led interventions and strict restrictions on youth access. However, the study suggests that in place of outright prohibition on ENDS, structured and informed discussions about ENDS and alternatives to smoking could be more effective in guiding youth toward harm reduction choices. Research by Arrazola et al. (2015) supports this approach and suggests that youth who were provided with open and factual dialogues about alternatives to smoking make better informed decisions about the use of tobacco ^[20]^. The present study also found that students were less likely to smoke. It is crucial that schools implement tobacco curriculum that includes understanding both the risks of smoking and the role ENDS plays in cessation, to ensure youth have an evidence-based perspective.

The relationship between knowledge, attitude, and behavior in this study supports the notion that knowledge influences perceptions, attitudes play a more direct role in determining smoking behavior. Consistent with research by Willett et al. (2019) that suggests youth who viewed ENDS as a harm reduction tool were more likely to use ENDS instead of cigarettes ^[4]^. Although, concerns regarding youth ENDS use remain valid, the results of this study suggest that ENDS uptake could still be optimized with strict regulatory frameworks rather than outright bans that would limit use for cessation while limiting initiation among non-smoking youth. Evidence-based regulation, rather than prohibition, is the sensible way forward, allowing access to ENDS for smokers while restricting any aggressive marketing behavior to non-smokers. Long-term outcomes of ENDS for cessation should be investigated in the context of youth behavior patterns and policy interventions.

## Study Limitations

Despite the comprehensive nature of this study, several limitations must be acknowledged. First, cross-sectional design limits the ability to establish causal relationships between knowledge, attitudes, socio-demographic factors, and ENDS use. Future studies should incorporate longitudinal research designs to examine behavioral changes over time. Second, the use of self-reported data introduces the possibility of social desirability bias, where participants may have underreported or overreported their ENDS use or attitudes toward vaping due to societal expectations. Another limitation is the lack of biochemical validation, such as cotinine testing, to confirm actual nicotine use, which may have resulted in misclassification of ENDS users and non-users. Additionally, convenience sampling was used, which, while ensuring accessibility, limits the generalizability of the findings to the broader youth population in Pakistan.

## Conclusion & Recommendations

The findings of this study revealed a complex relationship between knowledge, attitude, and smoking behavior among youth, indicating that knowledge alone is not a strong deterrent against tobacco use. Attitudes played a more significant role in predicting behavior, as Urban youth who viewed ENDS as less harmful or socially acceptable were more likely to use them, reinforcing the influence of peer acceptance and marketing strategies. Additionally, males with lower knowledge levels were more likely to engage in tobacco use, emphasizing the role of sociocultural influences. Furthermore, key predictors of smoking included financial independence, and behavioral risk factors such as alcohol and substance use, while protective factors such as parental monitoring and being a student reduced tobacco use. Furthermore, the study suggests that rather than viewing ENDS as a gateway to smoking, they may serve as a transitional aid for smokers aiming to quit. However, effective policies are needed to regulate access, ensuring that ENDS remain accessible to smokers while restricting aggressive marketing tactics that target non-smokers. A balanced approach that integrates harm reduction strategies, public education, and regulatory measures will be essential in shaping responsible tobacco control policies that prioritize both smoking cessation and youth prevention.

## Data Availability

The data underlying the findings of this study are not publicly available due to privacy and confidentiality agreements with the participants. However, the data can be made available upon reasonable request by contacting the corresponding author. All requests will be reviewed to ensure compliance with ethical and legal considerations.

## References

1. McNeill A, Brose L, Calder R, Bauld L, Robson D. Vaping in England: an evidence update including mental health and pregnancy, March 2020. Public Health England: London, UK. 2020.

2. Cullen KA. Notes from the field: use of electronic cigarettes and any tobacco product among middle and high school students—United States, 2011–2018. MMWR Morbidity and mortality weekly report. 2018;67.

3. Huang L-L, Baker HM, Meernik C, Ranney LM, Richardson A, Goldstein AO. Impact of non-menthol flavours in tobacco products on perceptions and use among youth, young adults and adults: a systematic review. Tobacco Control. 2017;26(6):709–19.

4. Willett JG, Bennett M, Hair EC, Xiao H, Greenberg MS, Harvey E, et al. Recognition, use and perceptions of JUUL among youth and young adults. Tobacco control. 2019;28(1):115–6.

5. Leventhal AM, Goldenson NI, Cho J, Kirkpatrick MG, McConnell RS, Stone MD, et al. Flavored e-cigarette use and progression of vaping in adolescents. Pediatrics. 2019;144(5).

6. WHO. Global Youth Tobacco Survey (GYTS): A comprehensive methodology for monitoring youth tobacco use’. 2021.

7. Islam MS, Saif-Ur-Rahman K, Bulbul MMI, Singh D. Prevalence and factors associated with tobacco use among men in India: findings from a nationally representative data. Environmental Health and Preventive Medicine. 2020;25:1–14.

8. Hameed A, Malik D. Public health practitioners’ knowledge towards nicotine and other cigarette components on various human diseases in Pakistan: a contribution to smoking cessation policies. BioMed Research International. 2022;2022(1):7909212.

9. Siddiqi K, Husain S, Vidyasagaran A, Readshaw A, Mishu MP, Sheikh A. Global burden of disease due to smokeless tobacco consumption in adults: an updated analysis of data from 127 countries. BMC medicine. 2020;18:1–22.

10. Qureshi FM, Bari SF, Zehra S, Mumtaz S. Rising trend of vaping products use amongst university students of urban setting in Pakistan: a cross-sectional survey. International Journal of Community Medicine and Public Health. 2025;12(1):112.

11. Mantey DS, Omega-Njemnobi O, Montgomery L. Flavored tobacco use is associated with dual and poly tobacco use among adolescents. Addictive behaviors. 2019;93:269–73.

12. Kowitt SD, Goldstein AO, Sutfin EL, Osman A, Meernik C, Heck C, et al. Adolescents’ first tobacco products: Associations with current multiple tobacco product use. PloS one. 2019;14(5):e0217244.

13. Alves RF. Health on you programme: development and implementation of web-based health education intervention for university students. Health Education Journal. 2022;81(6):667–78.

14. Jha P. The hazards of smoking and the benefits of cessation: a critical summation of the epidemiological evidence in high-income countries. elife. 2020;9:e49979.

15. Choi K, Forster J. Characteristics associated with awareness, perceptions, and use of electronic nicotine delivery systems among young US Midwestern adults. American journal of public health. 2013;103(3):556–61.

16. Wang TW. E-cigarette use among middle and high school students—United States, 2020. MMWR Morbidity and Mortality Weekly Report. 2020;69.

17. van Mourik D-JA, Nagelhout GE, van den Putte B, Hummel K, Willemsen MC, de Vries H. Did e-cigarette users notice the New European Union’s e-cigarette legislation? Findings from the 2015–2017 International Tobacco Control (ITC) Netherlands Survey. International Journal of Environmental Research and Public Health. 2019;16(16):2917.

18. Jackson A, Green B, Erythropel HC, Kong G, Cavallo DA, Eid T, et al. Influence of menthol and green apple e-liquids containing different nicotine concentrations among youth e-cigarette users. Experimental and clinical psychopharmacology. 2021;29(4):355.

19. Wills TA, Sargent JD, Knight R, Pagano I, Gibbons FX. E-cigarette use and willingness to smoke: a sample of adolescent non-smokers. Tobacco control. 2016;25(e1):e52–e9.

20. Arrazola RA, Singh T, Corey CG, Husten CG, Neff LJ, Apelberg BJ, et al. Tobacco use among middle and high school students—United States, 2011–2014. MMWR Morb Mortal Wkly Rep. 2015;64(14):381–5.

21. Audrain-McGovern J, Rodriguez D, Cuevas J, Sass J. Initial insight into why physical activity may help prevent adolescent smoking uptake. Drug and alcohol dependence. 2013;132(3):471–8.

22. Seo D-C, Kwon E, Lee S, Seo J. Using susceptibility measures to prospectively predict ever use of electronic cigarettes among adolescents. Preventive Medicine. 2020;130:105896.

23. Leventhal AM, Goldenson NI, Barrington-Trimis JL, Pang RD, Kirkpatrick MG. Effects of non-tobacco flavors and nicotine on e-cigarette product appeal among young adult never, former, and current smokers. Drug and alcohol dependence. 2019;203:99–106.

